# Tumor Regression Following Engineered Polyomavirus-Specific T Cell Therapy in Immune Checkpoint Inhibitor-Refractory Merkel Cell Carcinoma

**DOI:** 10.1101/2024.07.01.24309780

**Authors:** Yuta Asano, Joshua Veatch, Megan McAfee, Jakob Bakhtiari, Bo Lee, Lauren Martin, Shihong Zhang, Francesco Mazziotta, Kelly G. Paulson, Thomas M. Schmitt, Ariunnaa Munkbhat, Cecilia Young, Brandon Seaton, Daniel Hunter, Nick Horst, Marcus Lindberg, Natalie Miller, Matt Stone, Jason Bielas, David Koelle, Valentin Voillet, Raphael Gottardo, Ted Gooley, Shannon Oda, Philip D. Greenberg, Paul Nghiem, Aude G. Chapuis

## Abstract

Although immune check-point inhibitors (CPIs) revolutionized treatment of Merkel cell carcinoma (MCC), patients with CPI-refractory MCC lack effective therapy. More than 80% of MCC express T-antigens encoded by Merkel cell polyomavirus, which is an ideal target for T-cell receptor (TCR)-based immunotherapy. However, MCC often repress HLA expression, requiring additional strategies to reverse the downregulation for allowing T cells to recognize their targets. We identified TCR_MCC1_ that recognizes a T-antigen epitope restricted to human leukocyte antigen (HLA)-A*02:01. Seven CPI-refractory metastatic MCC patients received CD4 and CD8 T cells transduced with TCR_MCC1_ (T_TCR-MCC1_) preceded either by lymphodepleting chemotherapy or an HLA-upregulating regimen (single-fraction radiation therapy (SFRT) or systemic interferon gamma (IFNγ)) with concurrent avelumab. Two patients who received preceding SFRT and IFNγ respectively experienced tumor regression. One experienced regression of 13/14 subcutaneous lesions with 1 ‘escape’ lesion and the other had delayed tumor regression in all lesions after initial progression. Although T_TCR-MCC1_ cells with an activated phenotype infiltrated tumors including the ‘escape’ lesion, all progressing lesions transcriptionally lacked HLA expression. While SFRT/IFNγ did not immediately upregulate tumor HLA expression, a secondary endogenous antigen-specific T cell infiltrate was detected in one of the regressing tumors and associated with HLA upregulation, indicating in situ immune responses have the potential to reverse HLA downregulation. Indeed, supplying a strong co-stimulatory signal via a CD200R-CD28 switch receptor allows T_TCR-MCC1_ cells to control HLA-downregulated MCC cells in a xenograft mouse model, upregulating HLA expression. Our results demonstrate the potential of TCR gene therapy for metastatic MCC and propose a next strategy for overcoming epigenetic downregulation of HLA in MCC.

## Introduction

Merkel cell carcinoma (MCC) is an aggressive neuroendocrine skin cancer. In two retrospective studies, chemotherapy alone resulted in progressive disease and death within nine months of therapy start.^1,2^ Although checkpoint inhibitors (CPIs) have transformed the therapeutic landscape such that now 26% of patients can survive for more than five years,^3,4^ more than 60% of patients either do not respond or experience progression after CPIs, which underscores the necessity for alternative therapeutic strategies.

As CPIs rely on the presence of tumor-specific T cells,^5^ supplementing exogenous MCC-specific T cells could constitute a strategy to overcome resistance to CPI. MCC provides a valuable model to explore such an approach as more than 80% of MCC cases are driven by the Merkel cell polyomavirus (MCPyV).^6^ MCPyV oncoproteins large-T and small-T antigens (LTAg/sTAg) are integrated in the MCC cell genome and tumor cells require their expression for survival and growth.^7,8^ the unique expression in tumor cells reduces potential for on-target, off-tissue toxicity.^9^ MCPyV antigens are naturally immunogenic with several CD4 and CD8 T cells recognizing MCPyV-derived epitopes identified in MCPyV^+^ MCC patients.^10–12^ Among them, the presence of tumor infiltration of T cells specific for KLLEIAPNC (LTAg_15-23_), an epitope from a domain shared by LTAg and sTAg, correlates with improved survival.^13^ LTAg_15-23_ is restricted by human leukocyte antigen (HLA)-A*02:01, the most frequent HLA-A allele in the United States population, present in 34.6-49.7% of individuals depending on population groups.^14^

We previously targeted MCPyV^+^ MCC by adoptively transferring expanded autologous, non-transgenic LTAg_15-23_-specific CD8 T cells combined with avelumab which achieved a complete response in one CPI-naïve MCC patient.^15^ Although these results suggest that transferred MCPyV-specific T cells are associated with reduction of MCC burden, it is unknown whether they are also effective in a CPI-refractory setting. Moreover, as frequencies and functional avidities of MCPyV-specific T cells are highly diverse leading to inconsistent T cell products among patients,^13^ engineering T cells to express a high-avidity TCR could overcome some challenges encountered and allow the adoptive transfer of consistent numbers of functional TCR-T cells.

A major obstacle for T cell-mediated therapies for MCC is that tumor cells frequently downregulate HLA surface expression which can lead to escape from T cell recognition.^16,17^ Before CPIs became first-line therapy for MCC, more than 80% of MCCs were found to express reduced or undetectable HLA.^16,17^ We previously documented two patients whose tumors had intact HLA expression before receiving autologous tumor-specific T cells but subsequently lost expression of the targeted HLA, leading to MCC progression. As MCC-associated absence of HLA expression is typically mediated by transcriptional repression,^16,17^ this can potentially be re-upregulated by pharmacological interventions. Several approaches have shown promise against MCC cell lines in vitro: histone deacetylase (HDAC) inhibitors, low-dose single-fraction radiation therapy (SFRT), and cytokines such as interferon gamma (IFNγ) or beta (IFNβ).^16,17^ Both low-dose radiation and intralesional IFNβ have been used in patients with discrete and attainable MCC lesions.^16,18^ Systemic IFNγ was effective to force HLA expression sarcoma patients,^16,19^ but its efficacy in MCC patients is unknown.

Although chemotherapy agents such as etoposide have shown to force HLA expression in MCC cell lines,^16^ it is unknown how lymphodepleting chemotherapy (LDC) by fludarabine and cyclophosphamide (Flu/Cy) affects HLA expression on MCC cells. Flu/Cy increases the availability of the circulating homeostatic cytokines IL-7 and IL-15, which is critical for persistence and efficacy of chimeric antigen receptor (CAR)-T cells.^20^ It is yet unclear and of great interest whether TCR-T cell therapy used in the context of MCC specifically and solid tumors in general can be equally favorable and also force HLA expression.^21,22^

Here we identified an LTAg_15-23_-specific HLA-A*02:01-restricted TCR, denoted TCR_MCC1_. We engineered autologous CD4 and CD8 T cells with TCR_MCC1_ (T_TCR-MCC1_) for infusion into seven patients with metastatic MCPyV+ MCC in a first-in-human Phase-I clinical trial. To force HLA expression before T cell transfer or to prolong T cell survival, patients either received 8 Gy SFRT,^18^ systemic IFNγ or LDC immediately before infusions. Per standard of care, all infusions were followed by a CPI (avelumab). High-dimensional assays were applied to serial blood and tumor biopsies to examine the interaction of T_TCR-MCC1_ with MCC and correlate the observations to clinical outcomes.

## Results

### Identification of HLA-A*02:01-restricted MCPyV-specific TCRs from a patient who experienced MCC regression and HLA-matched healthy donors

To identify a TCR for adoptive transfer, we reasoned that we could isolate promising candidates from MCPyV-specific T cell clonotypes that localize in regressing tumor, as such TCRs would have adequate functional avidities to recognize and kill tumor cells.^23^ From our previous adoptive transfer studies with ex-vivo-expanded autologous T cells and a CPI, we identified one patient whose tumor regressed after receiving ex-vivo expanded autologous HLA-A*02:01-restricted LTAg_15-23_-specific CD8+ T cells (**Figure 1a**).^15^ High-throughput TCRL sequencing identified the top-10 most enriched clonotypes representing 89% of the infusion product (**Figure 1b, left**),^24^ among which six (TCR_R1-6_) preferentially accumulated in the regressing tumor (**Figure 1b, right**), indicating their likely role in mediating tumor regression. The corresponding TCRa chains were identified by single-cell VDJ sequencing (scVDJ-seq) of the same sample.

**Figure 1.**
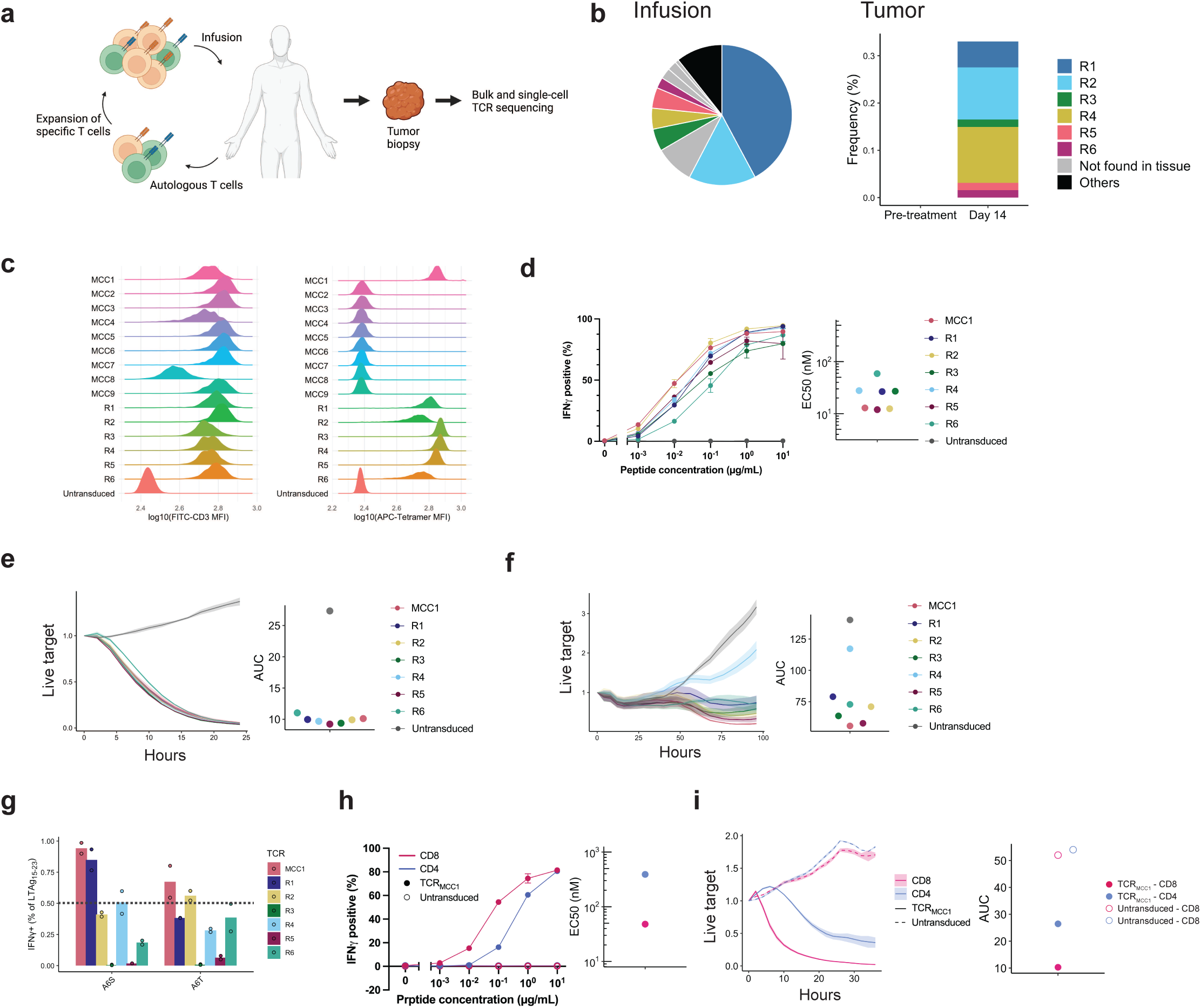
T cells transduced with the anti-MCPyV TCR identified from healthy donor PBMCs showed similar functional avidity but broader recognition of LTAg_15-23_ variants compared to those identified from responding patient-derived tumor-infiltrating T cells. **a** Schema of the expanded autologous T cell therapy with HLA-A*02:01-restricted LTAg_15-23_-specific CD8+ T cells resulting in MCC regression. **b** distribution in the infusion product (left) and frequency of the infusion-enriched clonotypes before and after T cell infusion in a regressing tumor biopsy (right). **c** CD3 expression (left) indicating surface expression of a functional TCR and LTAg_15-23_ p/HLA binding (right) by endogenous TCR knock-out Jurkat cells. **d** IFNγ production by CD8 T cells transduced with indicated TCRs exposed to TAP-deficient T2 B lymphoblastoid cell lines (T2-BLCL) exogenously loaded with indicated concentrations of LTAg_15-23_ (left) and mean half-maximal concentrations (EC50) (right) of three biological replicates for each TCR. 1 ng/mL LTAg_15-23_ equals 1 nM. **e, f** Growth kinetics of HLA-A2+ LTAg+ dermal fibroblasts (**e**) or WaGa cells (**f**) in the presence of CD8 T cells transduced with indicated TCRs. **g** IFNγ production by CD8 T cells transduced with indicated TCRs upon coculture with exogenously loaded LTAg_15-23_ variants (10 µg/mL). Horizontal line indicates 50% threshold below which variant recognition was impaired. Data from two biological replicates are shown. 50% **h, i** Same as (**d, e**) comparing CD8 vs. CD4 T cells transduced with TCR_MCC1_.

Although these TCRs constitute promising candidates, they might lack the highest-avidity clones, since chronic TCR stimulation by abundantly presented epitopes could delete T cells.^25^ While MCC patients often carry MCPyV-reactive memory T cells, healthy individuals lack such a population,^26^ indicating that their MCPyV-specific repertoire would consist of a naïve pool and retain high–avidity clones. Therefore, we expanded our search to HLA-matched healthy donors as previously described.^27^

We cloned the six patient-derived and nine healthy donor-derived TCRs into a lentiviral vector previously used clinically.^28^ To test their antigen specificity, we transduced them into TCR-deficient Jurkat cells. Since Jurkat cells lack CD8αβ coreceptor, their pHLA binding would indicate an ability to recognize the antigen when transduced in CD4 T cells. While all TCRs were expressed on the surface signifying functional TCRs (**Figure 1c, left**), one healthy donor-derived (MCC1) and all patient-derived (R1-R6) bound pHLA (**Figure 1c, right**).

### Healthy donor-derived TCR selected for clinical translation based on broader recognition of LTAg_15-23_ variants compared to patient-derived TCRs

We observed comparable avidities of CD8 T cells transduced with the healthy donor and patient-derived TCRs determined by responses of T cells to peptide-pulsed antigen presenting cells (EC50 11.9-58.8 nM) (**Figure 1d, left**). They all lysed HLA A*02:01-expressing human dermal fibroblasts transduced to express LTAg_1-278_ (**Fig. 1e**) as well as an MCPyV+ HLA-A2+ MCC cell line (WaGa)^29^ albeit with different kinetics (**Figure 1f**). However, only TCR_MCC1_ recognized both two clinically relevant LTAg_1-278_ variants—KLLEI**S**PNC and KLLEI**T**PNC—that occurs in 14% and 2% of cases respectively (**Fig. 1g**).^26^ Based on these results, we selected TCR_MCC1_ for clinical translation.

In addition to CD8 T cells, one can engage CD4 T cells through expression of a class-I-restricted TCR, thereby colocalizing engineered CD8+ and CD4+ T cells on the same tumor target to enhance therapeutic efficacy.^30^ Consistent with the observation that TCR_MCC1_ bound pHLA independent of CD8ab, CD4 T_TCR-MCC1_ produced IFNg in response to LTAg_15-23_ (**Fig. 1h**) albeit with a lower functional avidity (EC50 48.0 vs 390.0 nM). This translated into CD4 T_TCR-MCC1_ cells showing effective but less potent control of HLA-A2+ LTAg+ fibroblasts in vitro compared to their CD8 counterparts (AUC 10.3 vs 26.4) (**Fig. 1i**). Therefore, we planned to infuse both CD4 and CD8 T_TCR-MCC1_ cells, but based the cell dose on CD8 T_TCR-MCC1_ cell numbers only.

To probe the off-target recognition potential of T_TCR-MCC1_, we first assessed the contribution of each LTAg_15-23_ animo-acid residue to the reactivity of TCR_MCC1_. Substitutions in positions 4, 5 and 8 of LTAg_15-23_ to alanine abrogated T_TCR-MCC1_ IFNg production while substitutions in other positions were tolerated, signifying critical residues for TCR-p/HLA binding (**Supplementary figure 1a**). Next, a UniProt human proteome database^31^ search for peptides that shared amino-acids in positions 4, 5 and 8 with LTAg_15-23_ identified 31 peptides predicted to bind HLA-A*02:01 that had a three or four amino-acid difference from LTAg_15-23_ (**Supplementary Table 1**).^32^ One peptide derived from adenosine triphosphate-binding cassette subfamily G member 2 (ABCG2) (#22, ABCG2_331-339_) elicited T_TCR-MCC1_ reactivity (**Supplementary figure 1b**). However, HLA-A2+ fibroblasts transduced with the full-length of ABCG2 (**Supplementary figure 1c)** did not induce T_TCR-MCC1_ reactivity (**Supplementary figure 1d**), confirming the ABCG2 peptide is not endogenously presented. To probe T_TCR-MCC1_ potential for alloreactivity, T_TCR-MCC1_ were exposed to B lymphoblastoid cell lines (B-LCLs) expressing HLA allotypes covering a majority (HLA-A: 92%, HLA-B: 85%, HLA-C: 96%) of the European American population (**Supplementary Table 2, Supplementary figure 1e**).^33^ No reactivity was elicited confirming absence of potential for alloreactivity towards these hapotypes.^34^ Thus, TCR_MCC1_ was used to generate clinical-grade T_TCR-MCC1_ (see Methods).^28^

### Systemic IFNg as means to force HLA expression of HLA^low^ MCPyV+ MCC

MCC often downregulates HLA expression via epigenetic silencing rather than genomic deletion,^2^ permitting forced expression by therapeutic interventions. To identify systemic agents that could force MCC HLA expression and enable T_TCR-MCC1_ recognition, we compared clinically available IFNs (IFNβ (Avonex: IFNβ-1a, Betaseron: IFNβ-1b) and IFNγ-1b (Actimmune)) and HDAC inhibitors (valproic acid (Depakene), panobinostat (Farydak), and 5-azacytidine (Vidaza)) for their effect on WaGa cells. IFNγ was the most effective at forcing HLA expression of the WaGa cell line (**Supplementary figure 2a**) and the only agent that enable WaGa cells to stimulate CD8αβ+ mNeonGreen Jurkat reporter cells (**Supplementary figure 2b**). IFNg exposure at clinically achievable concentrations also forced HLA expression on four MCPyV^+^ MCC cell lines (Methods, **Supplementary figure 2d**). IFNg pre-treatment further enhanced lysis of the WaGa cell line by primary CD8 T_TCR-MCC1_ (**Supplementary figure 2c**). Based on these data, IFNg was used in combination with T_TCR-MCC1_ infusions in a subset of patients.

### T_TCR-MCC1_ preceded by forced HLA expression strategies and followed by CPIs induce tumor regression in two of seven CPI-resistant MCC patients without injuries to normal tissue

Seven patients (Pts) (NCT03747484) with CPI-resistant metastatic MCPyV+ MCC received a total of 10 T_TCR-MCC1_ infusions (**Table 1, Supplementary Figure 3**). Pts 1-4 received a first infusion of 1 x 1e8 CD8 (dose level 1) accompanied by 8.11 x 10e7-1.62 x 10e8 CD4 T_TCR-MCC1_. Pts 1, 3 and 4 received a second infusion of 9.38, 3.01 and 4.43 x 10e8 cells (dose level 2, target dose 10e9) respectively 112, 76 and 57 days after the first. Pts 5-7 received one infusion of 10e9 CD8 accompanied by 3.7-9.6 x 10e8 CD4 T_TCR-MCC1_. Pts received one of three interventions prior to T cell infusions: Pts 1, 2 and 5 received 8 Gy SFRT to non-index lesion(s) to trigger an abscopal effect five days prior to T_TCR-MCC1_ infusion.^35^ Pts 3 and 4 received standard lymphodepleting chemotherapy (LDC) of cyclophosphamide 300 mg/m^2^ and fludarabine 30 mg/m^2^ for three days ending 72 hours before infusions to enhance T_TCR-MCC1_ persistence. Pts 6 and 7 received systemic IFNγ1b (Actimmune 1,000,000 units/m^2^ 3 times week subcutaneously) a total of four weeks, starting five days before T cell infusions.^19^ Since LDC resulted in progression (discussed below), the intervention was discontinued and pt 5 received SFRT before Actimmune was approved for subsequent patients. All patients also received a standard dose/regimen of a CPI per standard of care (avelumab preferred, one year planned) starting 14 days after the first T_TCR-MCC1_ infusion.

**Table 1.** Information on the infusion products.

Although expected CPI-, chemotherapy- and radiation-associated side effects were observed, T_TCR-MCC1_ infusions were well tolerated with no occurrence of ≥ Grade 3 T_TCR-MCC1_-associated toxicities (**Table 2**).

**Table 2.** Patient background information and adverse events.

Among the seven pts who received T_TCR-MCC1_, one experienced tumor regression (Pt 2) and six experienced progressive disease within 28 days after infusions. However, amongst the latter, one experienced a delayed tumor regression (Pt 6, **Figure 2a**). Pt 2 had 14 sites of subcutaneous disease along the left leg (**Figure 2b**). Three sites received SFRT and regressed.

**Figure 2.**
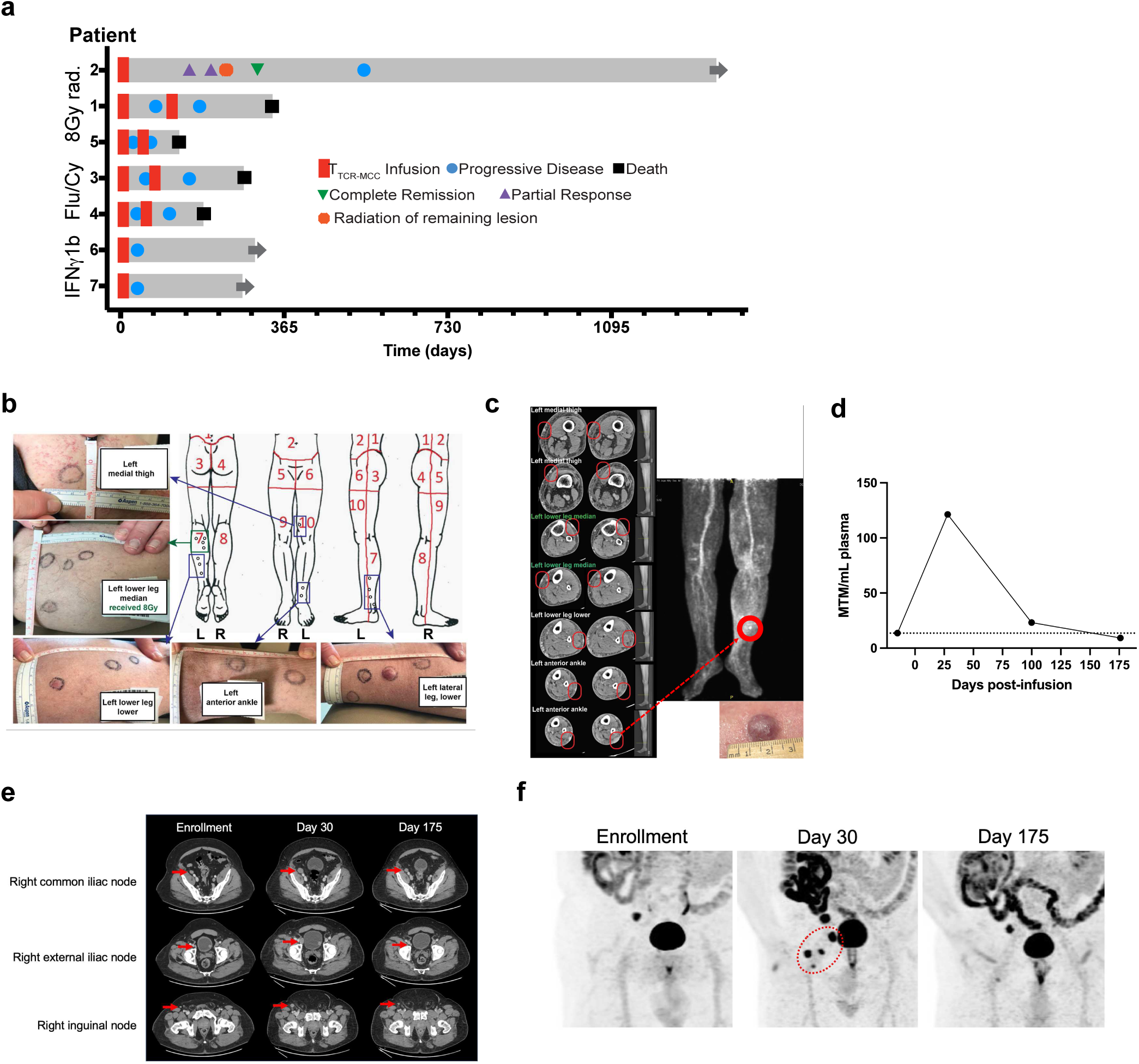
forced HLA expression, TCR_MCC1_, and CPIs induced tumor regression in 2/7 previously CPI-refractory metastatic MCC patients. **a** A swimmer’s plot showing clinical courses of the five enrolled patients. **b** Images and map of Pt 2 tumors at enrollment. **c** CT and PET scan images of Pt. 2’s remaining ‘escape’ lesion 105 days after TCR_MCC1_ infusion. **d-f** Pt 6 circulating tumor DNA (ctDNA) measurements (d), CT scan (e) and PET images (f) at indicated timepoints after TCR_MCC1_ infusion. Target lesions are indicated with red arrows and a circle.

Of the remaining 11 sites, 10 showed visible regression in the first month after T cell transfer. One “escape” lesion flattened but did not shrink (**Figure 2c**). received SFRT which resulted in the patient being disease-free until his systemic relapse 477 days after T_TCR-MCC1_ infusion.

Pt 6 had 3 metastases in the inguinal lymph nodes. After the start of the therapy, all the detectable metastases progressed over the course of the first two months, visualized by scans and cell-free tumor DNA (ctDNA) (**Figure 2d-f**). However, both scans and ctDNA showed regression in all inguinal lymph node metastases at three-month post-T cell infusion, down to a below-baseline level before progressing again.

### Circulating functional T_TCR-MCC1_ persist in CPI-resistant MCC patients

As expected from the higher functional avidity of TCR_MCC1-_trasduced CD8 compared to CD4 (**Fig. 1i**), higher frequency of CD8^+^ T_TCR-MCC1_ cells produced IFNg, TNFa and IL-2 upon cognate peptide recognition compared to CD4^+^ cells (**Figure 3a**). After infusion, the persistence of T_TCR-MCC1_ cells in circulation was monitored by measuring copy numbers of Woodchuck hepatitis virus posttranscriptional regulatory element (WPRE) in PBMCs (**Figure 3b**). Peak frequencies after DL1 and DL2 reached 3,460-93,227 and 3,132-631,431 copies/1e6 cells respectively between day 1 and day 7, except Pt 3 infusion 1 had a peak value of 574 copies/1e6 cells on day 48. Pt 3 and 4 who received LDC had lower peak WPRE copy numbers than the other patients (5.0e2–3.5e3 vs. 1.2e4–9.3e4 WPRE copies/1e6 cells for dose level 1, and 3.1e3–7.6e3 vs. 2.1e4–6.3e5 WPRE copies/1e6 cells for dose level 2). LDC increased homeostatic cytokine levels as expected reflected in a spike of serum IL-15 and IL-7 post-LDC (**Supplementary Figure 4**). We could detect cytokine production from both CD8+ and CD4+ T_TCR-MCC1_ cells after infusions also by flow cytometry (**Figure 3c**).

**Figure 3.**
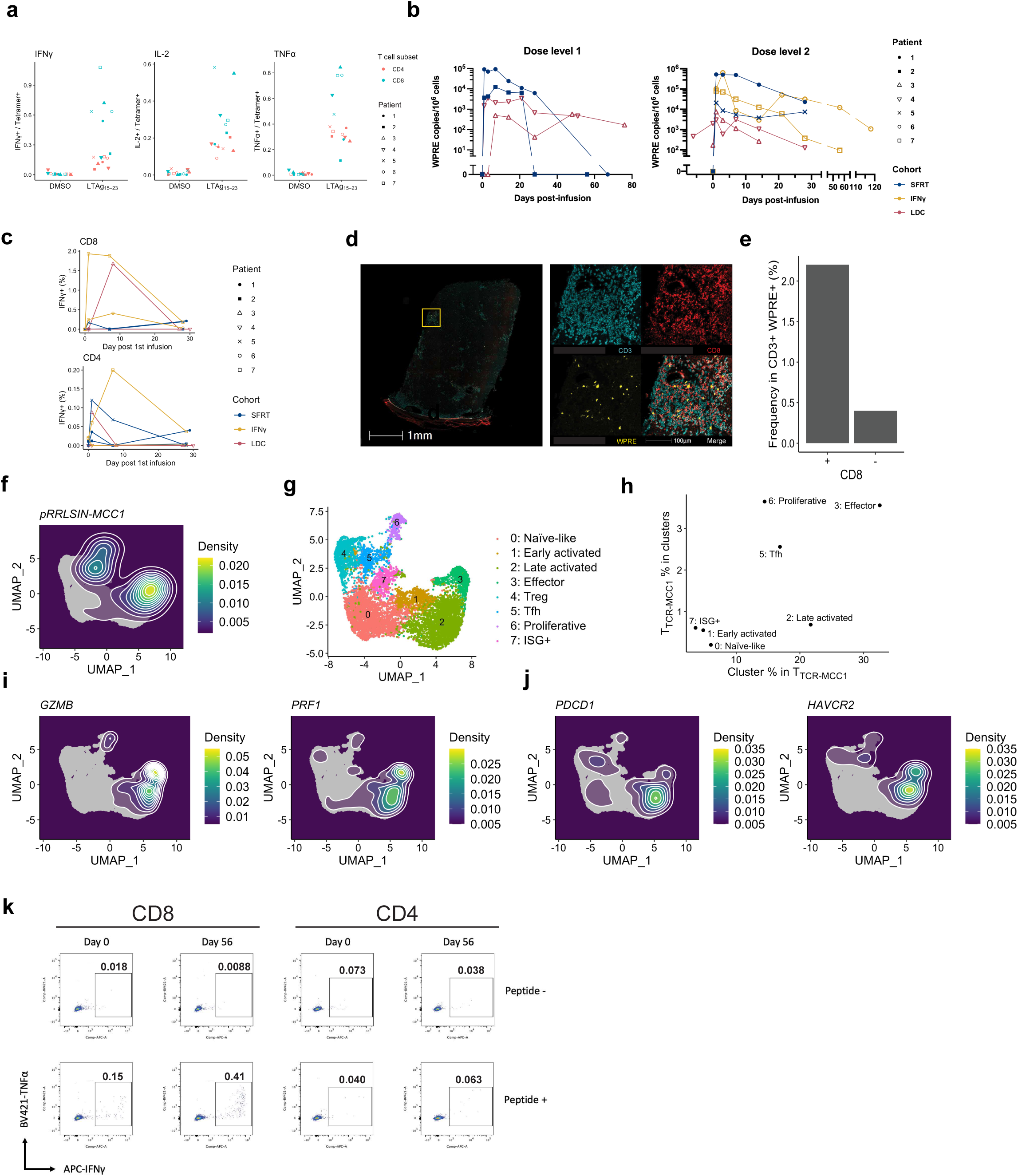
Functional T_TCR-MCC1_ cells persist in blood and preferentially infiltrate tumors. **a** Percent of CD4 and CD8 T_TCR-MCC1_ within the infusion products that produced IFNg, TNFa or IL-2 in response to LTAg_15-23_. **b** Circulating T_TCR-MCC1_ (WPRE copies/10^6^ cells) (y axis) at indicated timepoints (x axis) for all pts. **c** Percent of CD4 and CD8 T_TCR-MCC1_ that produced IFNg in response to 1µM LTAg_15-23_ at indicated timepoints. **d** CD3/CD8/WPRE (FISH) staining of Pt 2 ‘escape’ lesion. The right panel shows a zoomed-in image of the selected region in the left panel. **e** Quantification of CD8+ or CD8- CD3+ WPRE+ cells in (d). **f** Density of TCR_MCC1_ cells projected on the UMAP coordinate. **g** A UMAP plot of T cell populations with cluster annotations. **h** Frequency of T_TCR_-_MCC1_ cells in each cluster (y-axis) and frequency of each cluster in T_TCR_-_MCC1_ cells. **i, j** Density of cells that express indicated genes projected on the UMAP coordinate. **k** Cytokine production from expanded tumor-infiltrating T cells in Pt 6 biopsies.

### T_TCR-MCC1_ cells infiltrate MCC

Tumor biopsies were collected before, 14 days after each infusion then as clinically indicated (**Supplementary Table 3).** T_TCR-MCC1_ cells were detected by fluorescence in situ hybridization (FISH) probes matching WPRE in Pt 2’s ‘escape’ lesion (**Figure 2c**) and Pt 3’s progressive tumor obtained 56 days after the first infusion (**Figure 3d, Supplementary figure 5a**). In a single section of the Pt 2 day 105 tumor, we detected 6,721 CD3+ cells. Among them, 2.2% (148 cells) were WPRE+ CD8+, whereas 0.40% (27 cells) were WPRE+ CD8- (**Figure 3e**), indicating that CD8 T_TCR-MCC1_ cells preferentially infiltrated tumor compared with CD4.

To further characterize T_TCR-MCC1_ cells in tumors, we performed scRNA-seq on tumor biopsies. After analyzing each patient’s data separately (**Supplementary Table 4**), we integrated data that contained T_TCR-MCC1_ cells (Pt 2 day 105, Pt 3 infusion 1 day 56, Pt 6 day 14 and day 52, and Pt 7 day 10; **Figure 3f**, **Supplementary figure 5b**). Analysis of T cell populations found that 92.8% of T_TCR-MCC1_ cells (Pt 2: 16/17, Pt 3: 7/7, Pt 6: 50/54, Pt7: 4/5) expressed either *CD8A* or *CD8B*, which further demonstrates that CD8 T_TCR-MCC1_ preferentially infiltrated tumors compared with CD4. Within 9 T cell clusters we found (**Figure 3g, Supplementary figure 5c, Supplementary Table 5**), T_TCR-MCC1_ cells were most enriched in cluster 3 characterized by highly expressed genes associated with cytotoxic effector functions such as *GZMB* and *PRF1* (**Figure 3h and 3i, Supplementary figure 5d**).

Although T_TCR-MCC1_ cells expressed activation and cytotoxicity-associated genes, they were also identified in progressive lesions. Moreover, the distribution of T_TCR-MCC1_ cells on the UMAP embedding marginally overlapped with that of those expressing exhaustion marker genes, such as *PDCD1* (PD1) and *HAVCR2* (TIM3), indicating they could be terminally exhausted (**Figure 3j**). Therefore, we sought to test the functionality of tumor-infiltrating T_TCR-MCC1_ cells. To this end, we expanded T cells in pre-treatment and day 56 (progressive time point) tumor single cell suspension using CD3 stimulation and examined response to the peptide antigen. We detected higher frequency of cytokine-producing cells in day 56 (**Figure 3k**). We cannot attribute the cytokine-producing population entirely to T_TCR-MCC1_, since endogenous tumor-reactive T cells could have contributed as well. Nonetheless, the results indicate that functional tumor-reactive T cells were enriched in a post-treatment tumor.

### MCC HLA transcriptional expression correlates with tumor regression

Tumor progression despite the infiltration of apparently functional T_TCR-MCC1_ cells suggested that tumor cells employed an immune evasion mechanism. Since MCC often downregulate HLA, we probed tumor-associated HLA expression by IHC. Six of the seven treated patients who experienced progression within four weeks of infusion demonstrated low/absent HLA expression before T_TCR-MCC1_ infusions (**Figure 4a**). HLA also remained low/absent in five of those patients (Pts 1, 3-5, 7) after infusions. Tumor cells in Pt 2, the only patient who experienced MCC regression within four weeks of infusion, expressed HLA at baseline as well as in a regressing lesion 14 days after T_TCR-MCC1_ infusion. However, this patient’s “escape” lesion that remained progressive 105 days after transfer lacked HLA expression (**Fig. 4a**). Conversely, While Pt 6’s tumor had minimal HLA expression at the baseline and day 52 post-T cell infusion, tumor cells upregulated HLA at the time of delayed regression (day 118 post-transfer) correlating HLA expression with presence/absence of tumor regression post-T_TCR-MCC1_ infusion.

**Figure 4.**
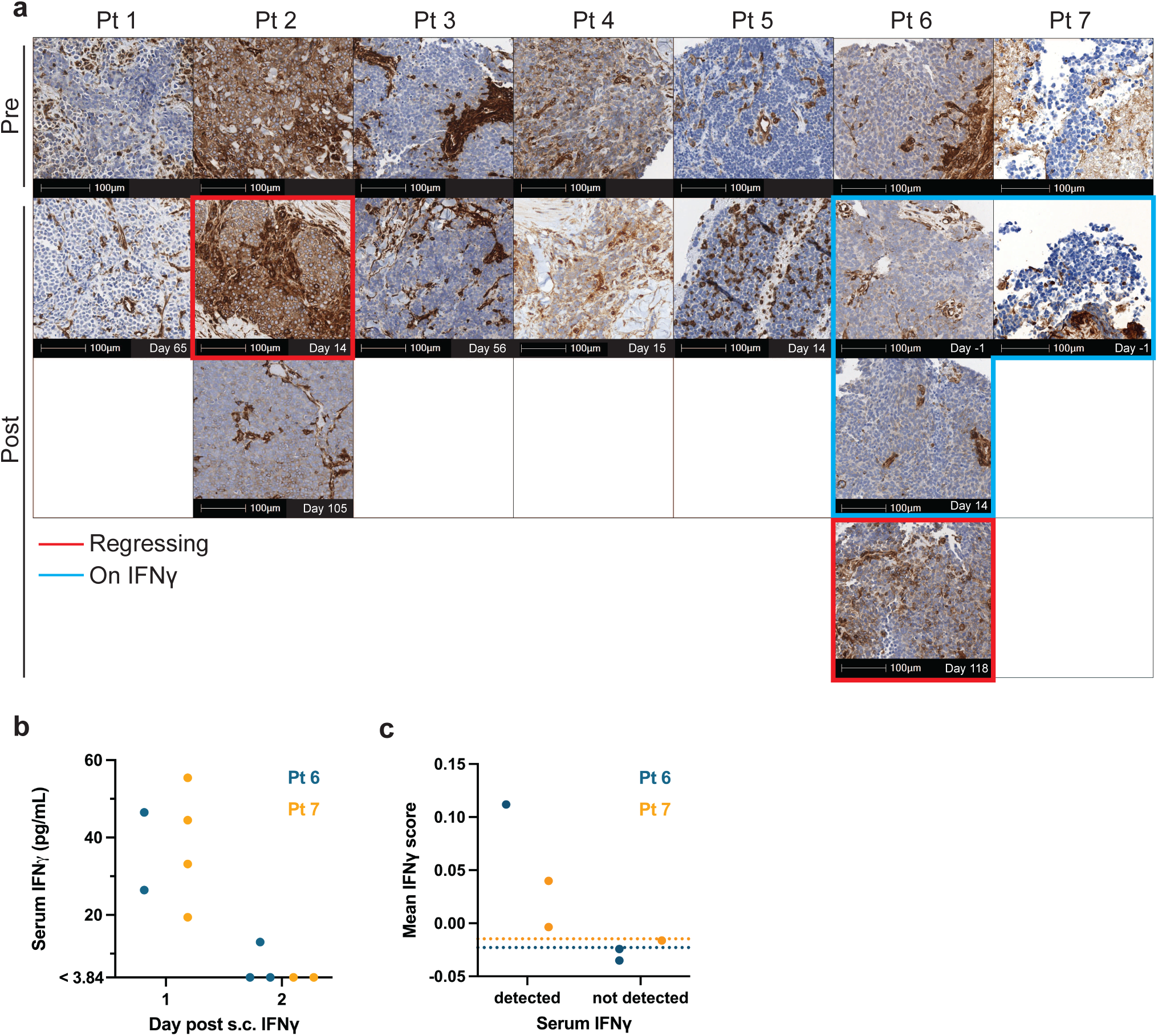
HLA expression/downregulation is associated with tumor regression/progression. **a** IHC of class-I HLA in tumor tissues. **b** Serum IFNγ level detected by Luminex in Pts 6 and 7 during the systemic IFNγ regimen. **c** IFNγ downstream gene scores calculated in Pts 6 and PBMC scRNA-seq data up to Day 28 post-T cell infusion. Each data point represents a single time point where scores of all cells were averaged. Dotted lines indicate basal levels at pre-IFNγ time points.

We performed whole exome sequencing on HLA-low/negative tumor samples of all patients (**Supplementary Table 6-9**), which revealed HLA-A*02:01 and *B2M* loci were intact in all patients. Pt 4 had a loss-of-heterozygosity event with deletion of one allele each of HLA-A/B/C but not HLA A*0201. These data indicate the lack of HLA expression was mediated by transcriptional repression rather than genetic deletion.

### SFRT, lymphodepletion and in particular IFNg, do not force immediate systemic MCC HLA expression sufficient to induce tumor regression

Of the patients who received SFRT, Pts 1 and 5 lacked baseline HLA expression and progressed. Although SFRT caused tumor regression of the radiated tumors which in turn could not be biopsied, other non-radiated distal lesions continued to lack HLA expression and show progression 65 and 14 days after T_TCR-MCC1_ infusion respectively (**Fig. 4a**), indicating circumscribed repercussions with no evidence of abscopal effect. We observed no HLA upregulation in Pts 3 and 4 who received LDC. MCC in these patients continued to progress 56 and 14 days after T_TCR-MCC1_ transfer respectively.

We obtained Pts 6 and 7 tumor biopsies four days after their first IFNγ administration. IHC did not detect HLA upregulation in these tissues (**Figure 4a**). S.c. IFNγ reached the systemic circulation, as we detected increase in serum IFNγ levels **(Figure 4b)**. However. the concentrations rapidly declined to below the detection limit (> 3.84 pg/mL) two days after administration (**Figure 4b**), reflecting the short systemic half-life of IFNγ.^36^ We detected downstream effects of IFNγ signaling in Pts 6 and 7 PBMCs by scRNA-seq only when IFNγ was detectable in sera, underpinning the transient activation of the IFNγ pathway (**Figure 4c, Supplementary Table 10**).

### Endogenous activated/expanded T and NK cells following T_TCR-MCC1_ infusion correlate with HLA expression and MCC regression

Given the short half-life of IFNγ and absence of its immediate effect in tumors, it is unlikely that administered IFNγ mediated HLA upregulation at the delayed regression in Pt 6. Furthermore, we could not detect T_TCR-MCC1_ cells in scRNA-seq of tumor biopsies on Day 118, indicating that endogenous immune cells could have driven the response.

To characterize immune cell population dynamics in Pt 6 tumor, we integrated scRNA-seq data of serial tumor biopsies obtained from a lymph node metastasis at five time points (Pre-IFNγ, Post-IFNγ/pre-infusion, Progressive: Day 14 and 52, and Regression: Day 118). Following low-resolution clustering (**Figure 5a, Supplementary figure 6a, Supplementary Table 11**), T and NK cell populations were re-clustered to resolve population complexity (**Figure 5b, Supplementary figure 6b, Supplementary Table 12**). This analysis identified eight major cell types, in which T and NK cells were divided into nine and two subpopulations respectively.

**Figure 5.**
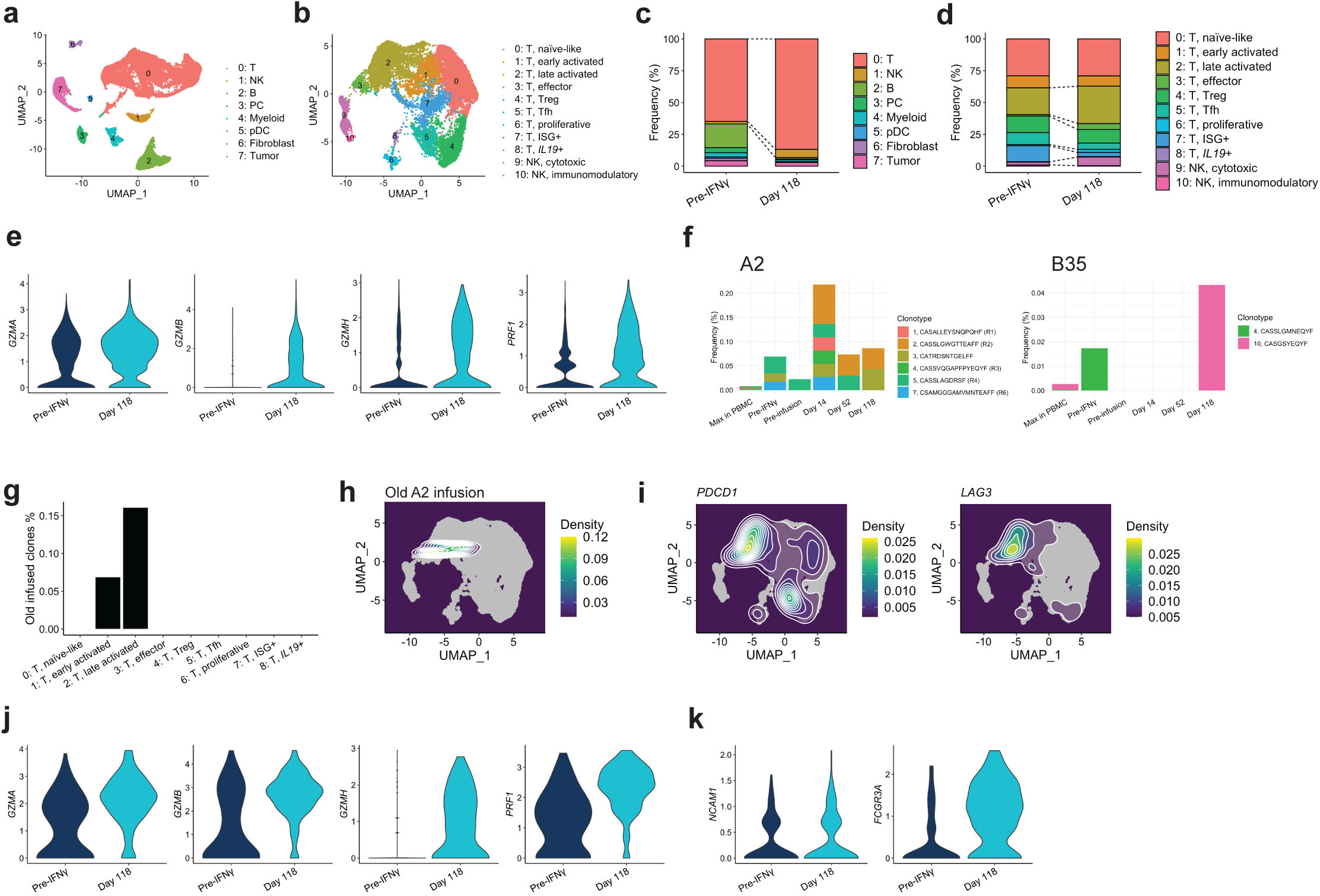
Activated endogenous T and NK cells expanded in a tumor showing delayed regression. **a, b** scRNA-seq cluster annotation of major cell types (a) and T/NK cell sub-clusters (b). **c, d** cluster frequency change from pre-treatment to Day 118, with (c, d) corresponding to (a, b) respectively. **e** Effector gene expression in T cells of sub-cluster 1-3 combined at the indicated time points. **f** TCR clonotypes in the infusion products for a previous endogenous T cell trial that were re-discovered in Pt 6 PBMC and tumors. “Max in PBMC” indicates the maximum frequency across tested PBMC time points. **g** Frequency of old HLA-A2-targeting infusion clones in T cell sub-clusters. **h** Distribution of the A2-targeting clones on the UMAP plot. **i** Density of cells that express indicated genes projected on the UMAP coordinate. **j, k** Effector (j) and phenotypic marker (k) gene expression in NK cells at the indicated time points.

By comparing cell type distributions in pre-treatment and day 118 tumors, we found that T cells expanded in the responding tumor (**Figure 5c**). Among them, Clusters 2 (late activated), 3 (effector), and 6 (proliferative) increase frequency within T cells, while Clusters 5 (Tfh) and 7 (type-I IFN-stimulated genes (ISG)+) decreased (**Figure 5d**), indicating T cells shifted toward more activated phenotypes. Indeed, when we compared gene expression in activated (Clusters 1-3) T cells on Day 118 vs. pre-treatment, they upregulated genes encoding effector molecules such as *GZMA*, *GZMB*, *GZMH*, *PRF1* (**Figure 5e, Supplementary Table 13**). These data suggest endogenous T cells had elevated cytotoxic potential at the regressing time point.

Pt 6 previously received autologous ex-vivo-expanded T cells targeting HLA-A*02:01-restricted LTAg_15-23_ and B*35:01-restricted sTAg_83-91_ (**Figure 1b, Supplementary figure 7a**). We detected some of the top-10 clonotypes enriched in the previous infusion products in tumor, up to three cells per clone in each time point (**Figure 5f**). Frequency in tumor was an order of maginitude higher than the maximum frequency in peripheral blood, showing their preferential localization in tumor. Clone 10 of the B35 infusion product was detected only on Day 118, although we could detect only one cell with this clonotype and none in quality-control-filtered gene expression data. Some of the endogenous LTAg_15-23_-specific (A2 infusion) clones were present in the gene expression data, where the majority had a late-activated phenotype expressing *PDCD1* and *LAG3* (**Figure 5g-i, Supplementary figure 7b and 7c**), consistent with their tumor specificity. Meanwhile, their phenotypic distribution contrasted with that of T_TCR-MCC1_ cells that were enriched in the effector-like phenotype, with several found in the proliferating population (**Figure 3h, Supplementary figure 7d and 7e**). This could reflect these old infusion clonotypes’ longer residency in tumor and/or better functionality, as less differentiated T cells have higher anti-tumor potential.^37^

In addition to T cells, NK cells—particularly those associated with higher cytotoxicity— expanded in the responding tumor (**Figure 5c and 5d**). NK cells also upregulated the genes encoding effector molecules on Day 118 (**Figure 5j, Supplementary Table 13)**. While *NCAM1* (CD56) level did not differ between pre-treatment and Day 118, NK cells in regressing tumor had higher expression of *FCGR3A* (CD16), associated with highly cytotoxic NK cells (**Figure 5k**).^38^ These results indicate that delayed activation of endogenous T and NK cells could have contributed to delayed tumor regression and HLA re-upregulation in tumor cells.

### MCC HLA expression and tumor control can be achieved by T_TCR-MCC1_ enhanced with CD8ab and the “Signal 2” CD200R/CD28 switch receptor

From our clinical trial, we identified two issues. First, CD4^+^ T_TCR-MCC1_ cells failed to infiltrate tumor compared with CD8^+^ cells. Engaging functional CD4^+^ T_TCR-MCC1_ cells could augment activity of CD8^+^ T_TCR-MCC1_ and other immune cells. Second, downregulated tumor HLA hindered T cell recognition which we could overcome with highly activated, cytokine-secreting antigen-specific T cells. To enhance persistence and function of CD4^+^ T_TCR-MCC1_, we modified the transgene by adding CD8αβ to support peptide-HLA recognition by CD4 T cells.^39^ Additionally, we employed the CD200R-CD28 immunomodulatory fusion protein to provide CD4 and CD8 T_TCR-MCC1_ cells with HLA-independent stimuli.^40^ CD200 is expressed in 84% of MCC,^41^ and all seven patients in our cohort (**Figure 6a**). We constructed a lentiviral vector incorporating TCR_MCC1_, CD8αβ, CD200R-CD28 connected with P2A and T2A (**Figure 6b**). To assess their function, we used a xenograft of naturally CD200+ WaGa cells in NOD-*scid* IL2Rg^null^ (NSG) mice.^42,43^ While T cells with TCR_MCC1_ alone did not reduce tumor burden, addition of CD8αβ and CD200R-CD28 allowed T_TCR-MCC1_ cells to control WaGa cells (**Figure 6c**). Both CD8 and CD4 T_TCR-MCC1_ cells with CD8αβ and CD200R-CD28 better infiltrated tumors and upregulated HLA on WaGa cells (**Figure 6d and 6e**). These results indicate that CD8αβ and CD200R-CD28 would be a promising strategy to enhance T_TCR-MCC1_ cell therapy against HLA-downregulated MCC.

**Figure 6.**
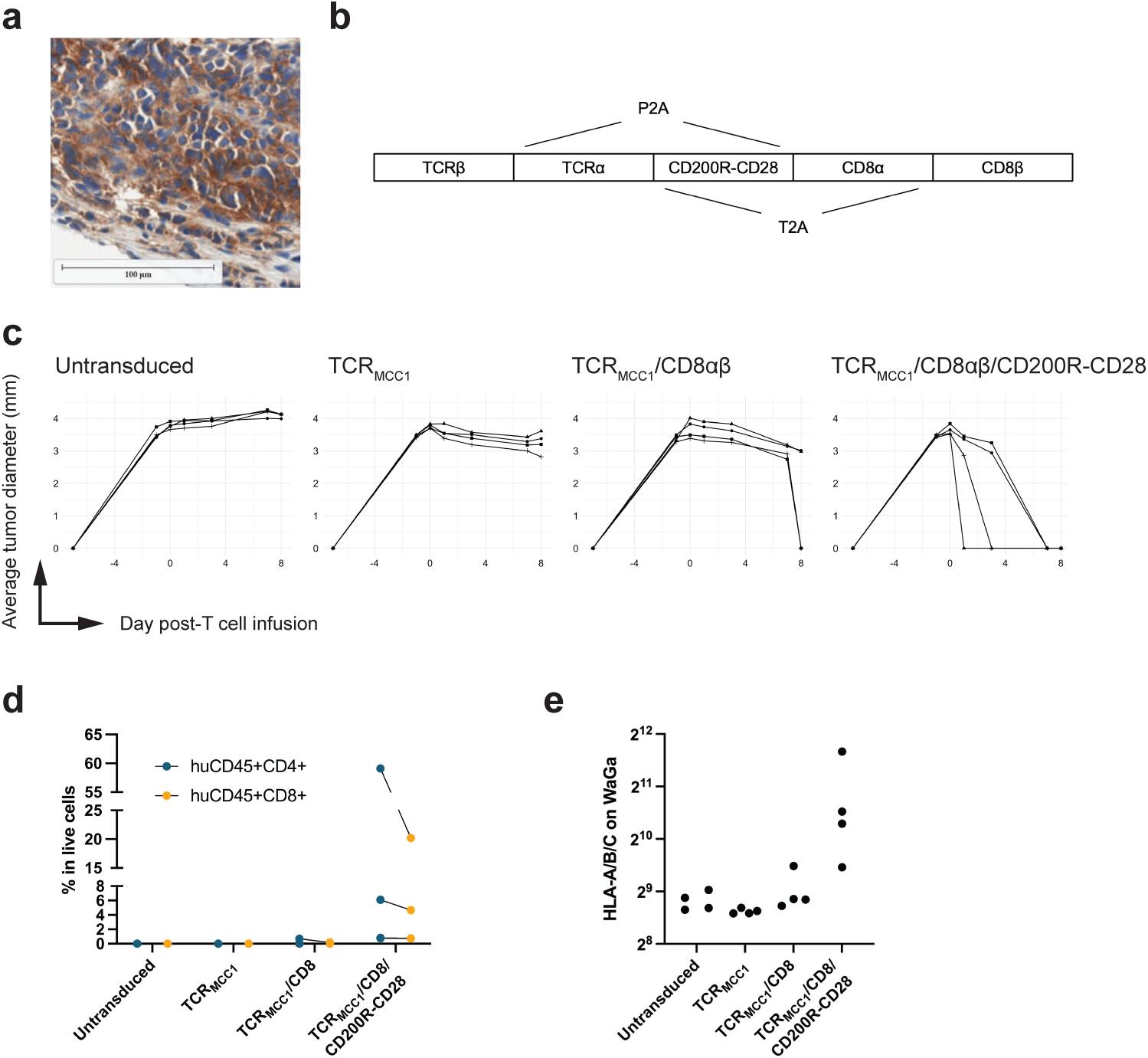
A CD200-targeting switch receptor allows TTCR-MCC1 cells control HLA-low MCC cells in mice. **a** CD200 expression in MCC. A representative image from Pt 1 is shown. **b** Design of TCR_MCC1_/CD200R-CD28/CD8αβ construct. **c** In vivo tumor control by T cells with the indicated transgene. 4 mice per arm. **d** Frequency of human CD4 and CD8 T cells in digested tumor single cell suspension. **e** HLA expression on tumor (huCD45- CD56+) cells measured by flow cytometry.

## Discussion

Here, we showed that MCPyV-targeting TCR-T cells can infiltrate tumors and induce tumor regression in CPI-refractory MCC patients. Occurrence of tumor regression was associated with class-I HLA expression in tumor cells. HLA was downregulated in all progressive lesions tested, with one patient showing de novo HLA downregulation, posing a common obstacle to the therapy. Meanwhile, another patient showed HLA re-upregulation in a lesion that showed delayed regression, demonstrating that immune response can reverse HLA loss in MCC.

As we observed in this and previous studies that HLA downregulation is common in MCC, we assessed SFRT and systemic IFNγ but we did not observe HLA-upregulating effects in vivo. Since we could not assess HLA expression in irradiated lesion that had regressed, we cannot exclude the possibility that SFRT did upregulate HLA locally without abscopal effects. A previous study that tested SFRT in MCC patients also noted the absence of abscopal effects in patients who showed response to the therapy,^18^ indicating that SFRT’s benefit might be locally limited for metastatic MCC patients. Subcutaneously administered IFNγ did have a systemic effect, as we observed upregulation of IFNγ-downstream genes in PBMC. Nevertheless, IFNγ did not induce sustained MCC HLA upregulation, presumably due to its short in vivo half-life. Prolonged exposure to IFNγ might also have triggered a negative feedback loop, limiting its efficacy.^44^ Another factor we need consider in using IFNγ is its potential immunosuppressive effects,^45^ which might have contributed to the disease progression we observed early after treatment in both patients who received IFNγ.

We used LDC in two patients to increase the expansion and persistence of transferred T cells, in part through the increase in homeostatic cytokines such as IL-7 and IL-15. While LDC increased those cytokines in circulation in the two patients, T_TCR-MCC1_ cells expanded poorly after each infusion compared with those who did not receive LDC. Moreover, tumor regression occurred rapidly but transiently after LDC. This antitumor effect would be a direct cytotoxic effect of Flu/Cy rather than transferred T cells, as MCC is highly sensitive to cytotoxic chemotherapy but responses are seldom durable.^46^ This cytotoxicity might have depleted available antigen or endogenous immune stimulatory cell populations, leading to inferior T cell persistence. In addition, LDC might also have impaired endogenous anti-tumor immune control such as tumor-specific T cells induced by previous CPI therapy. Such clearance of pre-existing anti-tumor immunity might underly faster disease progression that we observed in one patient following LDC. While LDC benefits persistence of adoptively transferred T cells in liquid tumor settings, its effect and the optimal dose in solid tumor settings are still unclear. A meta analysis of previous TCR-T cell therapies found higher-dose LDC was associated with toxicity but not objective responses.^21^ Another study of 11 solid tumor patients showed that LDC negatively affected the efficacy of neoantigen-specific T cell therapy.^22^ Although our cohort size is small, our results also suggest caution when using LDC with T cell therapy for solid tumor patients.

To enable TCR-T cells to recognize HLA-downregulated tumor cells, we used CD200R-CD28. Although CD200 is expressed on MCC cells, CD200 is expressed by other cells present in tumor microenvironments, such as B, regulatory T (Treg), follicular helper T (Tfh) and dendritic cells.^47^ It is interesting to test whether CD200-deficient tumor cells surrounded with CD200-positive tumor-infiltrating cells could confer better anti-tumor immunity by CD200R-CD28-equipped TCR-T cells. Moreover, CD200 is often expressed by non-MCC solid tumors of a neuroendocrine origine, such as small cell lung carcinoma.^41^ Testing efficacy of CD200R-CD28 against those non-MCC tumors would expand its clinical applicability.

Limitations of our study include the small cohort size and variability in T cell infusion products, such as dosage and transgene-positive cell frequency, though increasing dose levels from 1e8 to 1e9 did not appear to improve clinical outcome in our observation.

Our results demonstrate that TCR-T cell therapy targeting MCPyV is a promising treatment for CPI-refractory MCC patients, and further clinical investigation of enhanced TCR-T cell strategies are needed to provide new therapeutic options to patients.

## Materials and Methods

### Identification of ex vivo expanded infused HLA A*0201-restricted LTAg_15-23_-specific CD8 T cell clonotypes in a patient’s regressing tumor responding to a previous autologous T cell therapy trial

All clinical investigations were performed in compliance with the Declaration of Helsinki principles. The patient from whom the “patient TCRs” were isolated was enrolled on protocol #9245/NCT0258482 as well as the sample collection protocols #6585 and #1765 (biological sample collection) at the Fred Hutchinson Cancer Center (FHCC, Seattle, WA), all research activities were approved by the FHCC Institutional Review Board and the United States Food and Drug Administration. The patient provided a written informed consent. Briefly, this patient (9245-3) was a 59-year-old man with MCC that had initially presented as stage IIIB disease, metastatic at multiple sites. The patient had developed multiple relapses that had previously been treated with radiation; trial interventions were first systemic therapy.^15^ The patient received avelumab (anti-PD-L1) 10 mg/kg every two weeks and four infusions of MCPyV-specific T cells at dosages ranging between 0.8-3.9 x 10^9^ cells.^48^ Infused cells were of two specificities: HLA-B*35:01/STAg_83-91_ (FPWEEYGTL) and HLA-A*02:01/LTAg_15-23_ (KLLEIAPNC) epitopes, although only HLA-A*02:01 cells persisted in blood. The patient eventually experienced a complete remission and his regressing tumor was biopsied 14 days after the first infusion. To track infused clonotypes, DNA was extracted from tetramer-sorted infusion products and the day 14 tumor biopsy using a Qiagen DNeasy Blood and Tissue kit per manufacturers instruction. Samples were submitted to Adaptive Biotechnologies (Seattle, WA) for DNA amplification and sequencing of their TCRβ CDR3 sequences using a immunoSEQ hsTCRB Kit.^49^ Clonotypes were first identified in the infusion product and their frequency was tracked at the site of the regressing tumor.

### Cell culture for in vitro experiments

Human PBMCs (STEMCELL, #70500), primary T cells and DCs for TCR discovery experiments were cultured in X-VIVO15 media (Lonza) supplemented with 5% human serum and cytokines indicated below. Human PBMCs and T cells for in vitro functional assays were cultured in RPMI media (Thermofisher) supplemented with 5% human serum and 50 U/mL IL-2. 10 ng/mL IL-15 was also added at the secondary stimulation (detailed below). WaGa (a gift from Dr. Paul Nghiem, University of Washington), Jurkat (ATCC, #TIB-152), T2 (ATCC, #CRL-1992) and B-LCL cells (FHCC) were maintained in RPMI media supplemented with 10% Fetal Bovine Serum (FBS, Thermofisher). HEK293T (ATCC, #CRL-11268) and human dermal fibroblast cells (FHCC) were cultured in DMEM (Thermofisher) supplemented with 10% FBS. NK-92 cells were maintained in X-VIVO15 media supplemented with L-Glutamic acid (Thermofisher), 10% FBS and 100 U/mL IL-2. Media were replaced every two to three days.

### Construction of TCR-expression vectors

Identified TCRs were codon-optimized and synthesized by Genscript in an order of TCRβ-P2A-TCRα-WPRE. In order to promote inter-chain pairing of the transduced TCR, we introduced cysteine residues at amino acid positions 48 (Thr) of TCRα and 57 (Ser) of TCRβ constant regions respectively. We cloned the transgenes into pRRLSIN lentiviral vectors with a murine stem cell virus promoter digested with AscI (NEB) and SalI-HF (NEB) using NEBuilder HiFi DNA Assembly Master Mix (NEB) for further testing.

### Non-clinical lentivirus production

Hek293T cells were plated in 6-well plates at 5 x 10^5^ cells/well in 3 mL media. On the next day, cells were transfected with a mixture of plasmids encoding a transgene (150 ng/well) and a lentivirus backbone (molar ratio 1:2:4 = pMD2.G:pMDLg/pRRE:pRSV-Rev, 300 ng total/well) using Effectene reagent (Qiagen) following manufacturer’s protocol. Virus supernatant was collected after two days and concentrated with Lenti-X concentrator (Takara). Viruses were stored at -80 °C until being used.

### Production of TCR-deficient Jurkat cells and Nur77-mNeonGreen reporter cells

0.625 µL each of 160 µM clustered regularly interspersed short palindromic repeats (crispr) RNAs targeting TRAC (AGAGTCTCTCAGCTGGTACA, IDT) and TRBC1/2 (GGAGAATGACGAGTGGACCC, IDT) were duplexed with 1.25 µL of tracrRNAs (IDT) in duplex buffer (IDT) for 30 minutes at 37 °C. The duplexes were incubated with Cas9 proteins (61 µM, 1 µL, IDT) and polyglutamic acid (100 mg/mL, 1 µL) for 15 minutes at 37 °C. 4 µL of the mixture was added to 17 µL of P3 buffer (Lonza) and used to resuspend 1 x 10^6^ Jurkat cells. 20 µL of cell suspension was electroporated with 4D-Nucleofector X Unit (Lonza) with the EH-115 program. After one week of culture, CD3 negative cells were flow-sorted. To create Nur77 reporter cells, CD3-KO Jurkat cells were further modified to 1) knock out class-I HLA genes using CRISPR with a pan-class-I-HLA guide RNA (AAAAGGAGGGAGCTACTCTC), 2) overexpressing codon-optimized CD8B-P2A-CD8A (in the pRRSIN backbone) using lentiviral transduction and 3) knock in mNeonGreen in the NR4A1 (Nur77) locus using CRISPR/homology-directed repair (guide: ATGAAGATCTTGTCAATGAT).

### Donor T cell transduction and expansion

T cells were isolated from one-hour rested healthy donor PBMCs using EasySep Human T Cell Isolation Kit (STEMCELL), plated in a 12-well plate at 1 x 10^6^ cells/mL with 50 U/mL IL-2, and stimulated with 1:100-diluted T Cell TransAct (Miltenyi). One day later, lentivirus was added to the culture. On the next day, media were replaced. On day 4 after transduction, T cells were stained with 1:200 FITC-CD8, 1:200 PerCP/e710-CD4, and 1:400 APC-pMHC tetramer and sorted on an Aria II, separating CD4 and CD8 tetramer+ T cells. 2 x 10^5^ sorted T cells were mixed with 1 x 10^7^ mixed donor PBMCs (4000 rad irradiated) and 2 x 10^6^ B-LCL cells (8000 rad irradiated) with 50 U/mL IL-2, 10 ng/mL IL-15 and 30 ng/mL anti-CD3 antibody (clone: OKT3, Miltenyi) in 5 mL media in 6-well plates. T cells were expanded for 10 days with media replaced every two to three days and used for experiments or re-stimulated for another round before being used for co-culture assays.

### Intracellular cytokine staining

T2 cells were loaded with the wild-type LTAg_15-23_ peptide or variants A6S and A6T [KLLEISPNC and KLLEITPNC] at an indicated concentration for 1 hour, mixed with 1 x 10^5^ T cells at an indicated effector:target (E:T) ratio in a round-bottom 96-well plates in the presence of GolgiStop and GolgiPlug (1:1000, BD Biosciences). 16 hours later, cells were stained with anti-CD8, anti-CD4 and Live/Dead-Aqua (1:1000, ThermoFisher), fixed with BD Perm/Fix, washed with BD Perm/Wash, stained by 1:100 anti-IFNγ and analyzed on BD FACS Canto II.

### Fibroblast- and tumor-killing assays

Human dermal fibroblasts and WaGa cells were lentivirally transduced to express LTAg_1-278_-P2A-mCherry or mCherry respectively. 5 x 10^4^ T cells were co-cultured in 96-well flat-bottom culture plates (Corning) with 1 x 10^4^ target cells. Coculture with WaGa cells was supplemented with 50 U/mL human IL-2 (Peprotech). Killing was monitored every two hours with IncuCyte (Sartorius) by whole-well imaging with integrated fluorescence intensity as a readout.

### Self-/allo-reactivity testing of TCR-MCC1

**1. Alanin substitution of LTAg_15-23_**. LTAg_15-23_ variants having an alanine substitution at each amino acid position was synthesized (Genscript). T2 cells were loaded with those peptides and cocultured with CD8 TTCR-MCC1 cells to test IFNg production. **2. Search for self-peptides that share an amino acid sequence with LTAg_15-23_.** The UniProt human proteome database was searched for nine-mer peptides that 1) have E, I, and C at positions 4, 5 and 9, and 2) have at most a four amino-acid difference from LTAg_15-23_. This search found no hits with a one or two amino-acid difference from LTAg_15-23_, while three and 75 peptides that had a three or four amino-acid difference respectively. Within the 78 peptides, we selected the three peptides with a three amino-acid difference and 34 peptides with a four amino-acid difference that ranked higher than 5% in a prediction of binding to HLA-A*02:01 by NetMHCpan.^32^ Since UniProt human proteome contains unreviewed putative sequences, we filtered out peptides that were not found in basic local alignment search tool (BLAST) search.^8^ This retained 31 self-peptides as potential self-antigens for TCR-MCC1. We synthesized the 31 peptides (Genscript), loaded T2 cells with them and assessed TCR_MCC1_ reactivity using IFNγ production as a readout. 3. Testing alloreactivity to B-LCLs. B-LCL lines whose class-I HLA alleles cover the most frequent alleles in the US were obtained from the FHCC research cell bank. Individual LCL lines were co-cultured overnight with CD8 TTCR-MCC1 cells at a 5:1 E:T ratio to test IFNγ production.

### Clinical protocol including T_TCR-MCC1_ CD4 and CD8 T cells

The trial was approved by the FHCC Institutional Review Board, the US Food and Drug Administration, and registered at clinicaltrials.org as NCT03747484. All patients provided a written informed consent.

### Patient selection

Patients were enrolled with following selection criteria: HLA-A2 genotype confirmed by high-resolution typing. MCPyV-positive MCC was experimentally determined. Patients must have previously received a CPI for MCC and experienced no response, persistent disease or relapse after initially responding and detectable disease by RECIST 1.1^50^ was documented by imaging and/or clinical evaluation. Exclusion criteria included the presence of active uncontrolled auto-immune diseases, prior solid organ transplant, corticoid therapy at a dose equivalent of > 10mg of prednisone per day, concurrent use of other investigational agents or MCC-directed therapies, active uncontrolled infections and untreated brain metastasis. The clinical information for treated patients is summarized in **Table 1**. The sample size for this study was not based on formal power calculations, but on feasibility and the potential to provide descriptive information, determine whether further study was warranted and reveal if the toxicity was acceptable.

### Treatment Plan

Patients were eligible to undergo leukapheresis to collect T cells for transduction after they had met all inclusion and exclusion criteria. Pts 1,2, 3 and 5 received 8 Gy SFRT to a subset of detectable metastasis 3 to 5 day prior to T_TCR-MCC1_ infusions and Pts 4 and 5 received standard LDC of cyclophosphamide 200mg/m^2^ and fludarabine 30mg/m^2^ for 3 days, ending 72 hours before T_TCR-MCC1_ infusions. Pts 6 and 7 received 50 µg (1,000,000 U)/m^2^ IFNγ via a subcutaneous route three days per week for four weeks starting five days prior to T cell infusion. The first four patients received a first infusion aiming at 1 x 10^8^ CD8 T cells and accompanying CD4 T cells (dose level 1) and were planned to undergo a second infusion of 1 x 10^9^ CD8 T cells and accompanying CD4 T cells (dose level 2) as soon as 28 days later if they had < 5% of transgenic T cells of total CD8 T cells and progressive disease was documented. After safety had been confirmed, subsequent patients received the dose level 1 from the first infusion. Patients were monitored for toxicities, based on Common Terminology Criteria for Adverse Events v5.0.^51^ Core needle biopsies were obtained before, 14 days after each T_TCR-MCC1_ infusion and at the time of progression. Avelumab was started per standard treatment schedule 14 days after the first infusion and was planned to be pursued for 1 year.

### Clinical generation of T_TCR-MCC1_

All ex vivo manipulations involving processing of products destined for infusion were performed in the cGMP Cell Processing Facility (CPF) of FHCC. Substrate CD4 and CD8^+^ T cells for generating the TCR_MCC1_-transduced T cells (T_TCR-C4_) were obtained from an autologous PBMC donation (leukapheresis of patients enrolled in Protocol FHCRC #9845 Cells were stimulated with polymer-bound CD3 and CD28 (TransAct). On day +1 and day +2 of the stimulation, the responding cells were transduced using the pRRLSIN.cPPT.MSCV/TCR_MCC1_-P2A-TCR_MCC1_.wPRE lentiviral vector at a target multiplicity-of-infection of ≤ 3. Protamine sulfate (10 µg/mL), IL-2 (50 IU/mL), IL-21 (30 ng/mL), IL-7 (5 ng/mL), and IL-15 (1 ng/mL) were added, and the cells were centrifuged at 2,500 rpm for 90 minutes at 30 °C, and then incubated overnight at 37 °C. On day +12 (+/-2) were harvested for infusion. Products were freshly infused in 5 of 8 infusions. Alternatively, the cells were thawed and washed before infusion. Quality control of the infused products included assessment of inserted lentiviral vector copy number per cell (≤ 5 copies/cell), envelope VSV-G DNA by qPCR as a surrogate marker for RCL (< 10 copies/50 ng DNA), and binding to the LTAg_15-23_ HLA-A2-restricted tetramer (≥ 30% of live cells).

### Clinical sample collection for flow cytometry and sequencing experiments

**1. PBMCs.** PBMCs were enriched from whole blood patient samples using Ficoll-Hypaque. Whole blood was diluted 1:1 with Dulbecco phosphate buffered saline (DPBS), layered on top of 12 mL Ficoll-Hypaque in 50 mL conical tubes, and centrifuged for 20 minutes at 1,000 g with no brake. PBMCs were washed twice in DPBS (400 g, 10 minutes, low brake) and cryopreserved in FBS containing 10% dimethyl sulfoxide and stored in liquid nitrogen until being used. **2. Tumor biopsies.** Tumor needle biopsies were collected from patients and either formalin-fixed (for IHC), snap-frozed (spatial transcriptomics) or stored in ice-cold RPMI (for scRNA/VDJ-seq). Samples collected in were processed on the same day with a Tumor Dissociation Kit (Miltenyi) and gentleMACS C Tubes following a manufacturer’s protocol. Single cells were washed in DPBS supplemented with 0.04% bovine serum albumin (Sigma, #A1595) and cryopreserved in CryoStar CS10 media (STEMCELL) until samples were batched for further processing.

### Transgenic T cell tracking by WPRE qPCR

Genomic DNA (gDNA) were prepared from patient PBMC using Qiagen DNeasy Blood and Tissue kit. 500 ng gDNA per sample were amplified in 25 µL reactions using 2x Power SYBR Green master mix (Roche) and 0.5 µM each forward and reverse primers targeting WPRE or the *ALB* gene. The amplification and signal quantification were performed using LightCycler 480 II (Roche). Plasmids containing one copy of either WPRE (epHIV7) or ALB (hAlbumin_pCMV6-AC) were used as standards.

Thermal cycling condition:

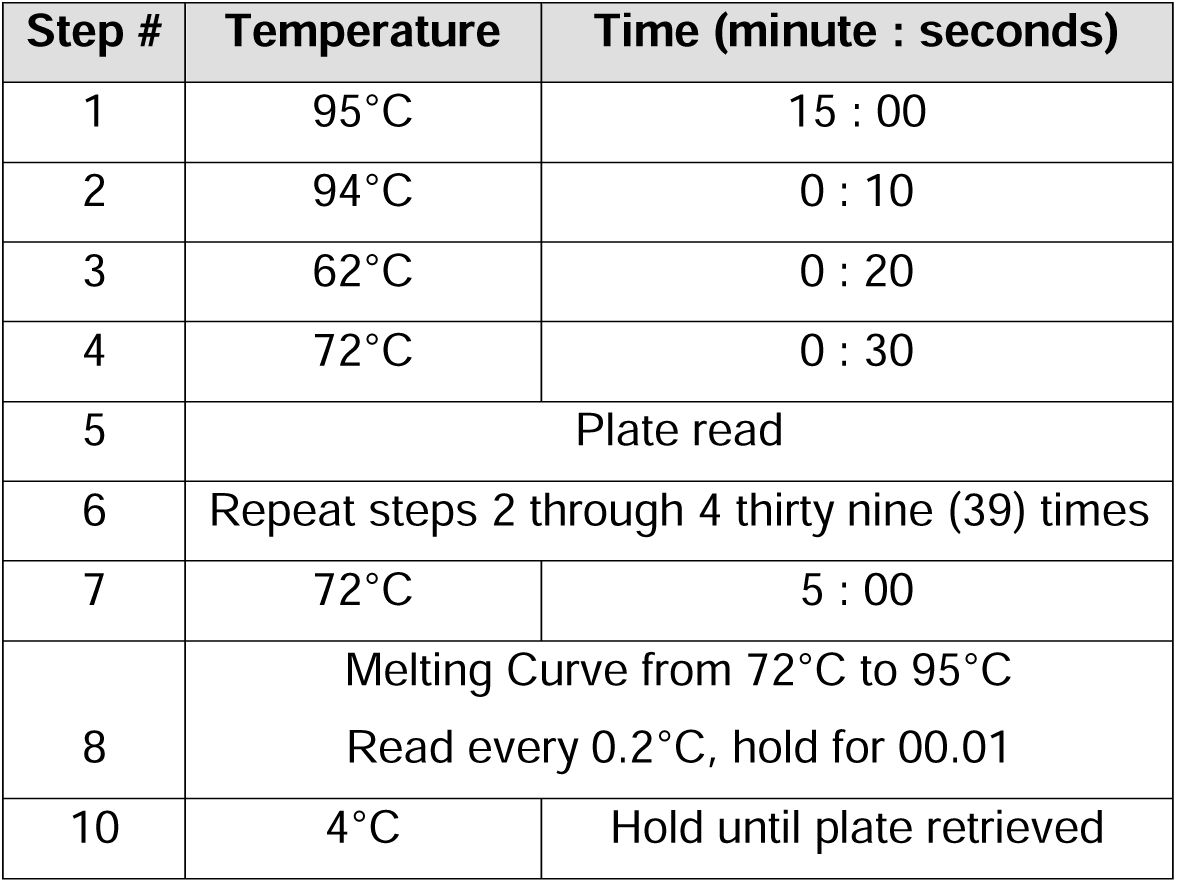

| Step # | Temperature | Time (minute : seconds) |
| --- | --- | --- |
| 1 | 95°C | 15 : 00 |
| 2 | 94°C | 0 : 10 |
| 3 | 62°C | 0 : 20 |
| 4 | 72°C | 0 : 30 |
| 5 | Plate read |  |
| 6 | Repeat steps 2 through 4 thirty nine (39) times |  |
| 7 | 72°C | 5 : 00 |
| 8 | Melting Curve from 72°C to 95°C<br>Read every 0.2°C, hold for 00.01 |  |
| 10 | 4°C | Hold until plate retrieved |

### Detection of KLL15-23-reactive cells in patients’ PBMCs

Cryopreserved patient PBMCs and infusion products were thawed and washed once in T cell culture media (RPMI + 5% human serum) and then resuspended in 2 mL of pre-warmed culture media to be rested overnight in an incubator at 37 °C 5% CO_2_. After resting overnight, cells were counted manually by hemocytometer, and then plated (∼5e5 cells per well) into 96 well plate format for peptide stimulation. LTAg_15-23_ was added at 10 µg/mL in combination with BD GolgiStop and GolgiPlug (1:1000). Cells were incubated for six hours and stained with 1:200 PE/Cy7 anti-CD19, FITC anti-CD8, PerCP/e710 anti-CD4 and 1:1000 Live/Dead-Aqua. Cells were fixed with BD Perm/Fix for 15 minutes at 4 °C, washed with BD Perm/Wash, and resuspended in BD Perm/Wash and left overnight at 4 °C. Internal stain was performed the next morning and cells were stained by 1:100 APC anti-IFNγ, PE anti-IL-2, and BV421 anti-TNFa for 15 minutes at 4 °C, cells were washed with 100 mL FACS buffer, resuspended in 150 mL FACS buffer and analyzed on BD Fortessa X-50.

### Pt 6 tumor-infiltrating T cell expansion and cytokine production assay

Cryopreserved digested tumor single cell suspension were thawed then washed once with T cell culture media (RPMI + 5% human serum) plus penicillin/streptomycin (Thermofisher). After washing, cells were immediately assembled into the rapid expansion protocol: 1e7 irradiated mixed PBMCs (three individual donor PBMCs from STEMCELL), 2e6 irradiated B-LCLs in combination with 50 U/mL IL-2 (Peprotech), 10 ng/mL IL-15 (Peprotech), and 1.5 mL anti-CD3 antibodies (OKT3) in 5 mL culture volume. Culture was expanded for 10 days, with culture media was replaced every 2 or 3 days, with the last media change being one day before functional testing and phenotyping. Cells were stimulated with LTAg_15-23_ as described for patients’ PBMCs.

### Serum cytokine Luminex assay

Cytokines were measured by Luminex multiplex assay. Samples and standards were diluted on a polypropylene dilution plate. Antibody-coupled microbeads were mixed by vortexing for 10 seconds to resuspend. Microbeads were combined by diluting stocks into wash buffer such that each bead population was present at approximately 2000 beads per 50 µL (∼ 1:150 dilution). 50 µL/well of diluted microbeads were added to all wells and washed on a 96 well magnetic plate with 140 µL of wash buffer (0.1% BSA, 0.05% Tween20 in PBS). 50 µL/well of standards and samples were added in duplicate and shaken on an orbital shaker at 500 rpm for at least 30 minutes in the dark, and then incubated overnight at 4 °C in the dark. The plate was washed twice as before using magnetic plate. 50 µL/well of combined biotinylated detection antibodies were added. Incubated for 60 minutes on an orbital shaker at 500 rpm in the dark. Detection antibodies were removed by washing twice. 50 µL/well of streptavidin:phycoerythrin (PE) conjugate (Prozyme # PJRS27) were diluted to 5 micrograms per milliliter in assay buffer and added to the wells. Incubated for 30 minutes on an orbital shaker at 500 rpm in the dark, followed by two washes. 100 µL/well wash buffer were added and shaken on an orbital shaker at 500 rpm for at least two minutes. PE intensity was measured immediately on Luminex 200 instrument. A five-parameter logistic standard curve is generated for each cytokine, with sample concentrations calculated from these curves.

### scRNA-seq

Sample preparation and library preparation was done following the user guide for Chromium Next GEM Single Cell 5’ Reagent Kits v2 (Dual Index) (CG00331 Rev D, 10X Genomics). For PBMC samples, cryopreserved cell suspension were thawed and washed once with PBS containing FBS. Digested tumor biopsies were washed once and subjected to Dead Cell Removal Kit (Miltenyi). Cells were counted, diluted and processed with Chromium Controller targeting 10,000 cells per sample. Constructed gene expression and TCR V(D)J libraries were quality-checked with Agilent 4200 Tapestation, followed by dual-index paired-end sequencing on Illumina NovaSeq S2 (gene expression) or NextSeq P2 (V(D)J). Sequencing reads were aligned to reference human genome GRCh38 provided by 10X Genomics using cellranger v6.1.1. For the alignment of gene expression reads, the TCR_MCC1_ and complete MCPyV sequences (NC_010277.2) were appended to the reference genome.

### scRNA-seq data analysis

Data analysis was done using Seurat package v4.1.0 on R v4.2.0. Poor quality cells were filtered out from gene-expression scRNA-seq data by the maximum mitochondrial read ratio and the minimum detected gene count as detailed in Supplementary Table 4. For tumor data, datasets with a median feature count below 300 were not used for analyses as poor-quality data. Data were analyzed and annotated for each patient separately, followed by integration and further analyses. For the analyses of T_TCR-MCC1_ cells (Figure 3), T cells from T_TCR-MCC1_ cell-containing dataset were extracted and integrated.

Data processing and differential expression analyses followed recommendations of Seurat v4 pipeline (https://satijalab.org/seurat/articles/archive). QC-filtered data were normalized with the SCTransform() v2 function. Cluster marker genes and pairwise differential gene expression (Pt 6, Day 118 vs. pre-IFNγ) were tested using FindAllMarkers () or FindMarkers() respectively (i.e. Wilcoxon rank-sum test) following the PrepSCTFindMarkers() function. P-values were corrected for multiple testing using the Bonferroni procedure. Heatmaps were generated using the top-10 marker genes (based on corrected p-values) for each cluster. Several genes were not included in the scale.data slot of integrated Seurat objects in the normalization and integration process. Such genes were scaled and added to the scale.data slot for the sole purpose of generating heatmaps. When scaled values were higher than 3 or lower than –3, they were cut off to 3 or –3 respectively to allow for plotting all genes within a single range of expression levels.

### IFN**γ**-downstream gene score calculation

PBMC scRNA-seq data from multiple time points were integrated for each patient separately. Expression level of IFNγ-downstream genes was scaled across cells and then averaged across genes to calculate IFNγ scores.

### Whole exome sequencing

Cryopreserved PBMCs and formarin-fixed paraffin-embedded (FFPE) tissue sections were processed by Personalis using ImmunoID NeXT platform.^52^

### Immunohistochemistry

Anti-HLA class I immunohistochemistry was performed using mouse mAb clone EMR8-5 from Abcam (catalog #ab70328). Following routine deparaffinization, FFPE sections undergo heat induced antigen retrieval in a pH 6.0 citrate-based buffer (Diva Buffer, Biocare Medical) for 20 minutes at 95 °C. Staining was performed on an automated intelliPath instrument (Biocare Medical) using an anti-mouse polymer conjugated to horse radish peroxidase (Leica Biosystems). DAB (3,3′-Diaminobenzidine) chromogen (Biocare Medical) was used to create a brown pigment visible using standard brightfield microscopy.

For WPRE FISH and CD3/CD8 staining, FFPE tissues were sectioned at four µm onto positive-charged slides and baked for 1 hour at 60°C. The slides were loaded to the Leica Bond Rx Autostainer platform (Leica, Buffalo Grove, IL) to start the run. The slides were baked and dewaxed using Leica Bond reagents for dewaxing (Dewax Solution). Antigen retrieval was performed at 95°C for 20 minutes using Leica Epitope Retrieval Solution 2 followed with 0.5% Triton-X/PBS solution pretreatment at 40 °C for 60 minutes. After all the pretreatment steps, the slides were blocked with hydrogen peroxide for 10 minutes and then incubated with ACD Target probe, WPRE (ACD #517728), Positive control probe Human PPIB (ACD #313908) or Negative control probe DapB (ACD #312038) at 42°C for 120 minutes. After the probe incubation, the staining continues with the RNAscope 2.5 LS Reagent Kit-Brown (ACD #322100) for the amplification and detection steps, followed by Opal fluor 570 (Akoya Biosciences #OP-001003) at 1:1000 for 10 minutes. To perform CD3 and CD8 antibody staining, the stripping steps were performed at 100°C for 20 mins using Leica Epitope Retrieval Solution 2. Endogenous peroxidase was blocked with 3% H2O2 for 5 minutes followed by protein blocking with TCT buffer (0.05M Tris, 0.15M NaCl, 0.25% Casein, 0.1% Tween 20, pH 7.6 +/- 0.1) for 10 minutes. A CD3 (Thermo #RM9107, 1:1000 dilution) primary antibody was applied for 60 minutes followed by the OPAL Polymer HRP Ms+Rb (by Akoya Biosciences #ARJ100EA) for 20 minutes. A tertiary tyramide signal amplification (TSA)-amplification reagent Opal fluor 520 (Akoya Biosciences #OP-001001) was added at 1:100 for 20 minutes. After the excess HRP was quenched, CD8 staining was performed with an anti-CD8 primary antibody (Agilent #M710301-2, 1:800 dilution) and a tertiary TSA-amplification reagent Opal fluor 650 (Akoya Biosciences #OP-011005). Slides were removed from the Bond Autostainer and stained with DAPI (10ug/ml distilled water) for 5 minutes, rinsed in water, and coverslipped using Prolong Gold Antifade reagent (Invitrogen #P36930). The slides were cured for 24 hours at room temperature and the images of the slides were acquired on the Vectra Polaris Imaging System. Images were analyzed using the HALO software (Indica Labs).

### Mouse xenograft model

For monitoring tumor control, 1e7 WaGa cells were engrafted to the left flank of 8-12-week-old female NSG mice (maintained at and sourced from Fred Hutch animal facility). Seven days later, 1e7 T cells (1:1 mixture of CD4:CD8) were injected into mice via the tail vain. Tumor diameter was measured with a caliper three times a week. For analysis of infiltrating immune cells and tumor HLA expression, T cells were injected 14 days after tumor engraftment, and tumors were collected after another 14 days. Tumors were chopped into small pieces and digested into single cell suspension in DMEM media containing 2 mg/mL collagenase-D (Sigma) and 20 µg/mL DNase-I (Sigma) for 30 minutes at 37 °C using gentleMACS Dissociator (Miltenyi) and gentleMACS C Tubes (Miltenyi). Single cells were stained with fluorophore-conjugated antibodies and analyzed on LSRFortessa (BD).

## Data Availability

All data produced in the present study are available upon reasonable request to the authors

## Funding

BlueBird Bio, Inc.

Affini-T Therapeutics, Inc.

National Institute of Health (P01CA225517)

## Acknowledgements

This research was supported by NIH P01 grant (grant ID: P01CA225517), Bluebird Bio and Affini-T Therapeutics. YA thanks Japan Society for the Promotion of Science for a research fellowship. Authors thank shared resources (Flow Cytometry, Experimental Histopathology, Immune Monitoring, Preclinical Modeling and Fred Hutch Innovation Lab) at Fred Hutch for their through supports.

## Author contributions

AGC and KGP conceived the study. AGC, YA, JV wrote the manuscript. JV, KGP supervised clinical studies. YA, MM designed and performed experiments. YA analyzed data. JB, LM, BL, AM, DH, BS, MS assisted and/or performed experiments. SZ, FM, CY, TS, DK, PDG, PN provided expertise and feedback.

## Supplementary

**Supplementary Figure 1.**
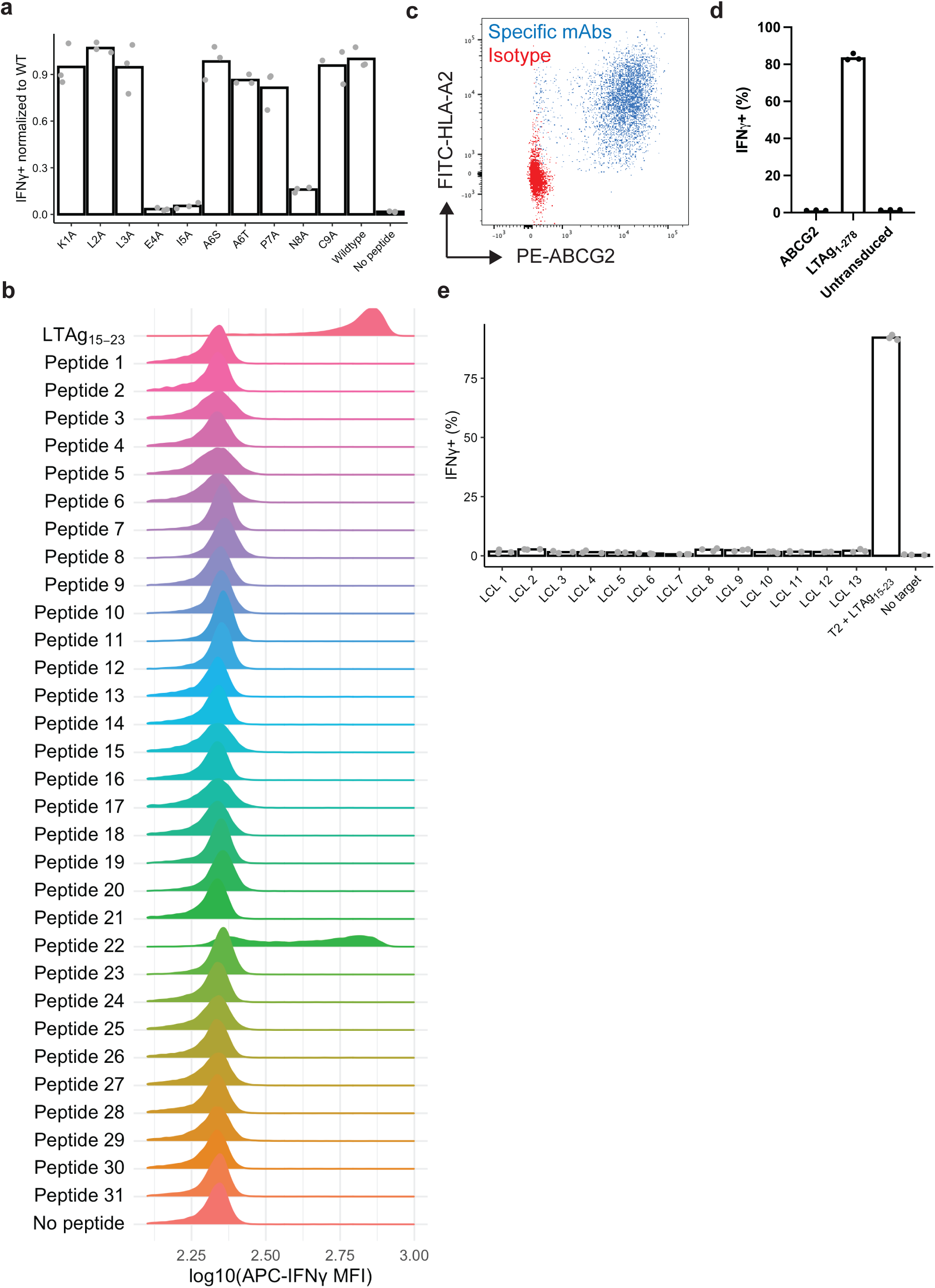
TCR_MCC1_ is neither self-nor alloreactive. **a** IFNγ production of CD8 T_TCR-MCC1_ cells upon coculture with T2 cells loaded with alanine mutants of LTAg_15-23_. Positive cell frequency was normalized by the mean values of unmutated LTAg_15-23_. **b** IFNγ production of CD8 T_TCR-MCC1_ cells upon coculture with T2 cells loaded with indicated self-peptides that has a similar sequence to that of LTAg_15-23_. LTAg_15-23_ was used as a positive control. **c** Dermal fibroblasts expressing HLA-A2 and ABCG2. **d** IFNγ production of CD8 T_TCR-MCC1_ cells upon coculture with indicated human dermal fibroblasts. **e** IFNγ production of CD8 T_TCR-MCC1_ cells upon coculture with indicated LCLs. LTAg_15-23_-loaded T2 cells was used as a positive control.

**Supplementary Figure 2.**
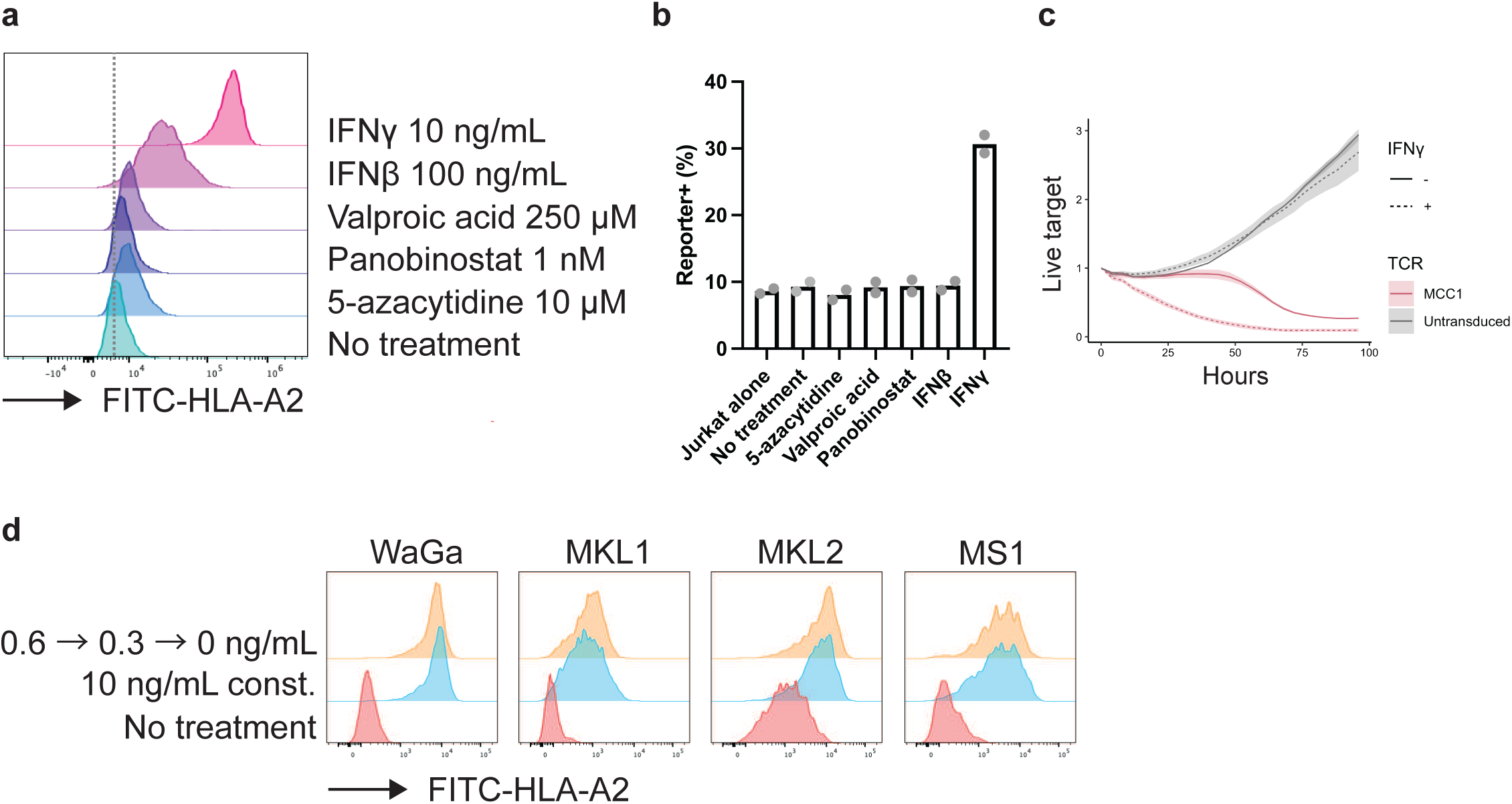
IFNg upregulates HLA on MCC cell lines. **a** HLA-A2 expression on WaGa cells measured after a 3-day culture with the indicated reagents. **b** NeonGreen expression frequency in Jurkat reporter cells after overnight coculture with WaGa cells pre-treated as in (a). **c** In vitro coculture assay using TCR_MCC1_-transduced primary CD8+ T cells against WaGa cells with or without 3-day pre-treatment with 10 ng/mL IFNg. **d** HLA-A2 expression on MCC cell lined with the indicated pre-treatment.

**Supplementary Figure 3.**
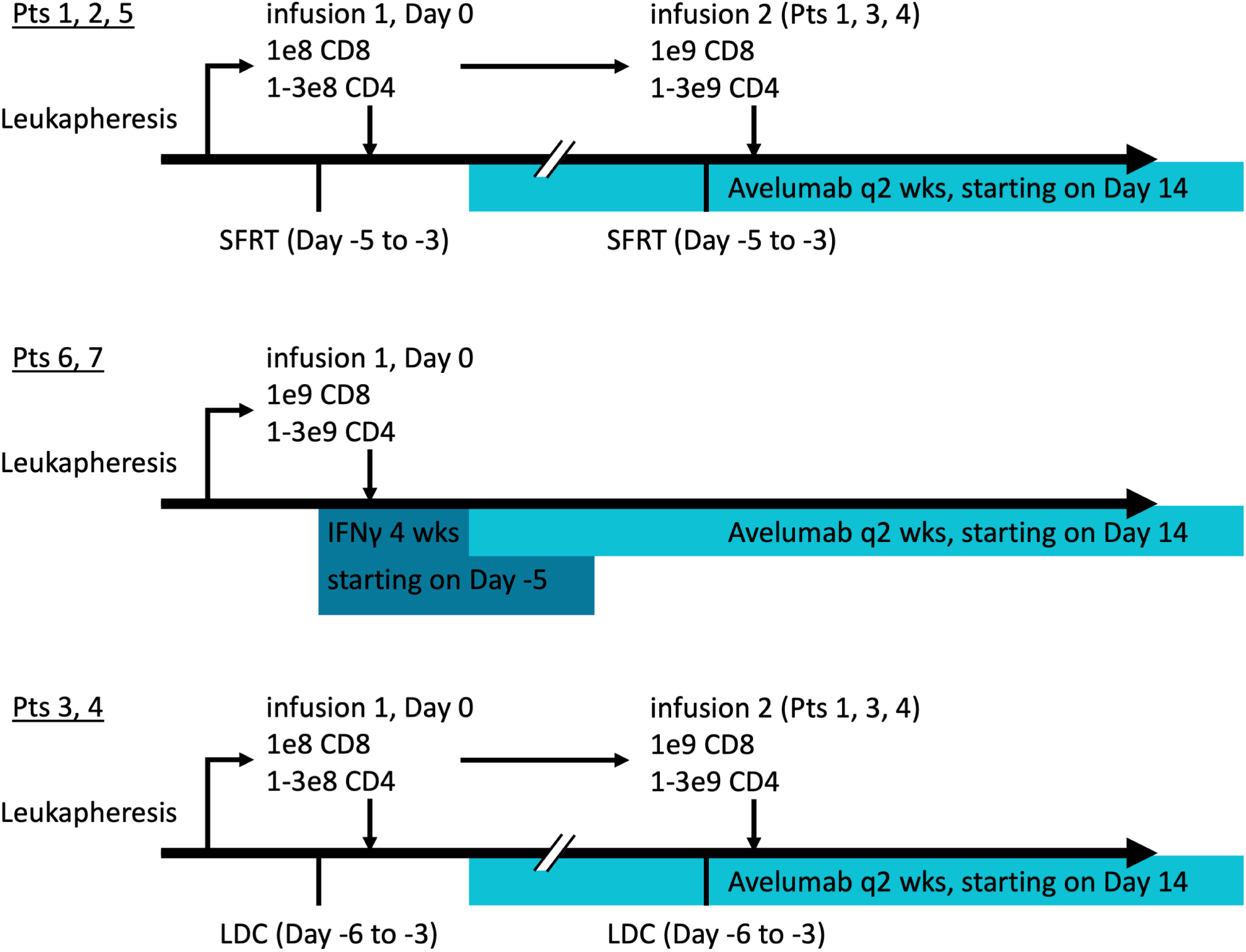
Clinical trial scheme.

**Supplementary Figure 4.**
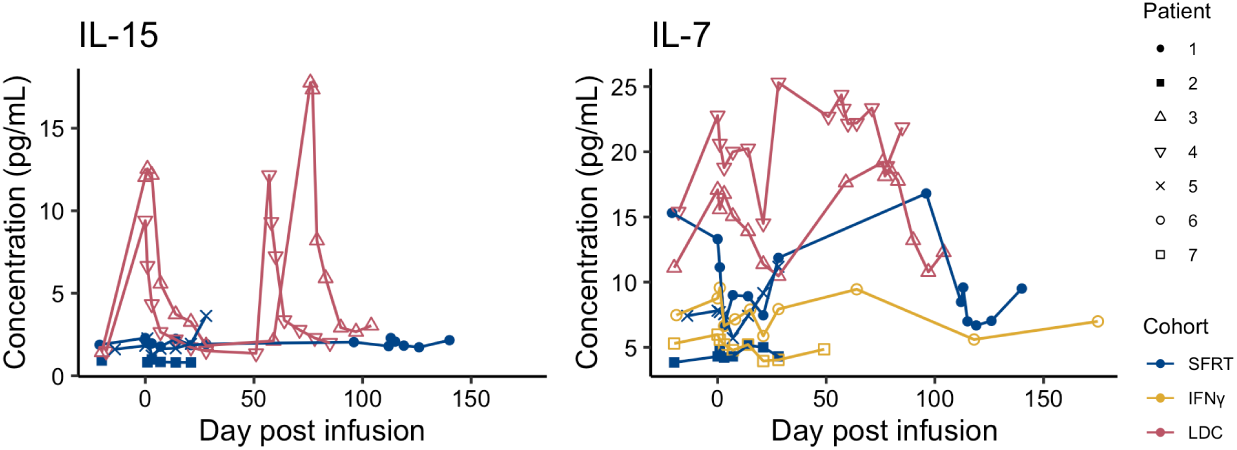
LDC increased serum concentrations of IL-15 and IL-7.

**Supplementary Figure 5.**
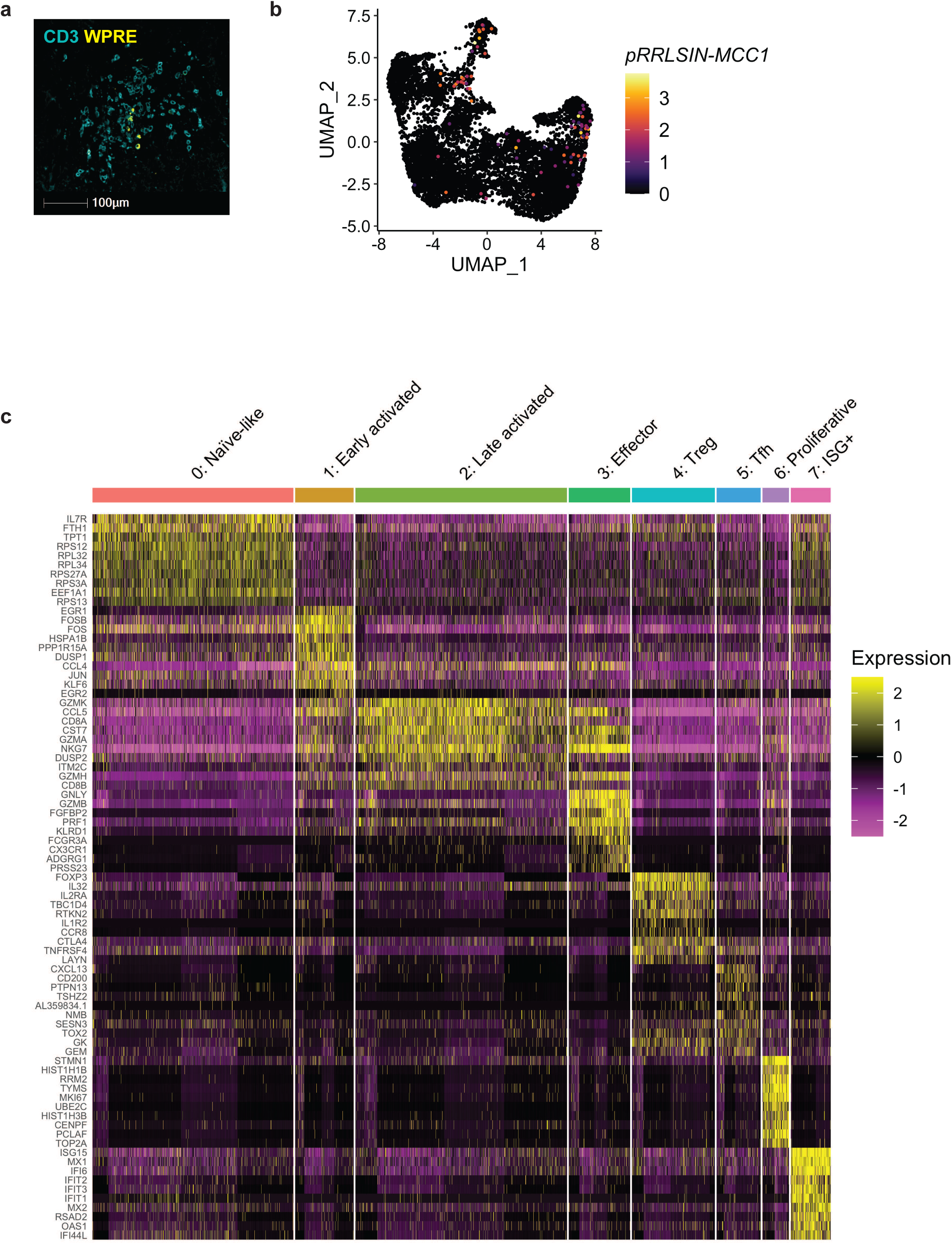
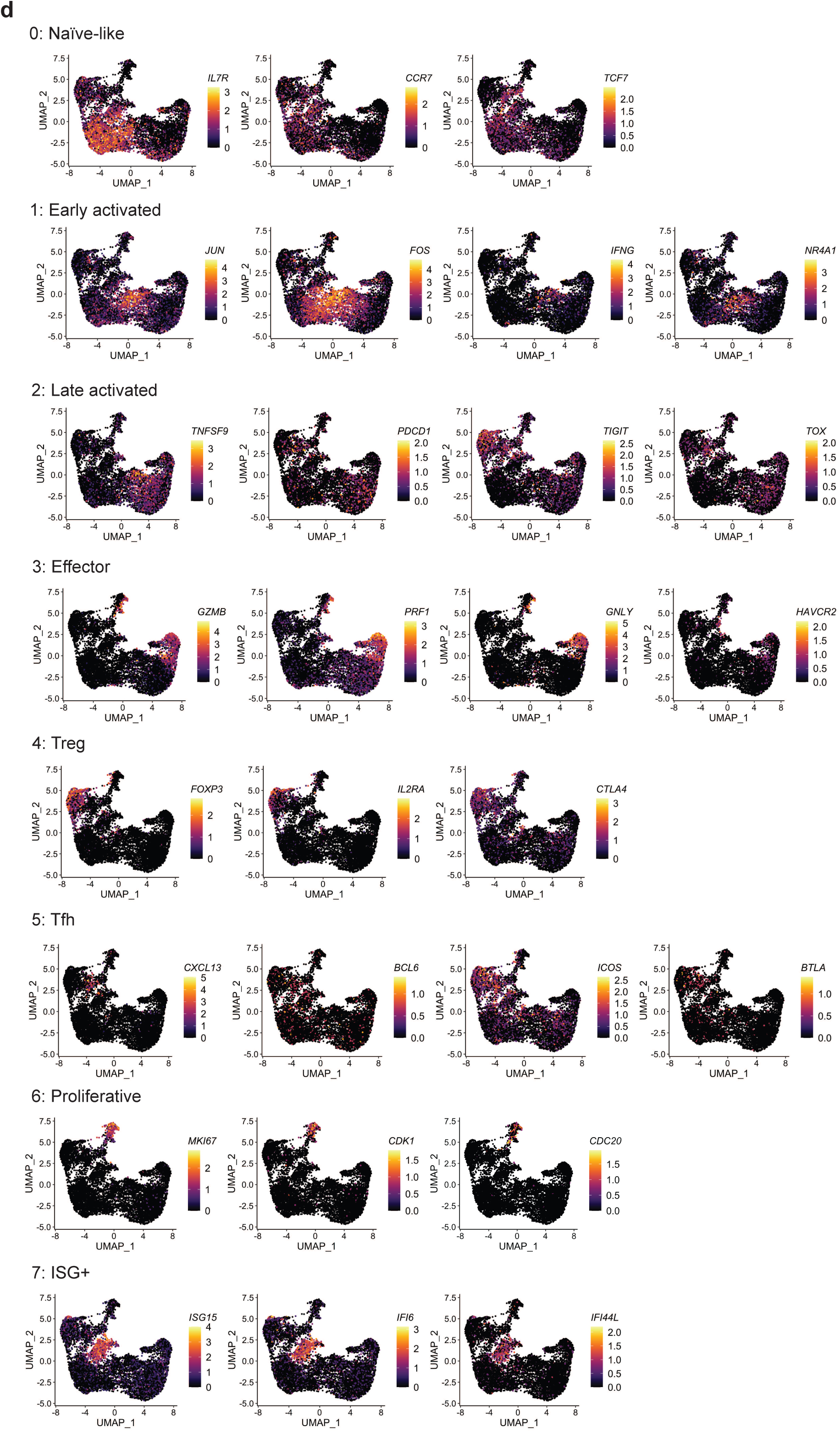
T_TCR-MCC1_ infiltration and T cell phenotypes in tumor. **a** CD3/WPRE IHC in Pt 3 infusion 1 day 56 tumor. **b** TCR_MCC1_ transcript expression on the UMAP embedding. **c** Heatmap showing expression of top-10 marker genes for T cell sub-clusters. **d** Representative marker genes expression for T cell sub-clusters on the UMAP embedding.

**Supplementary Figure 6.**
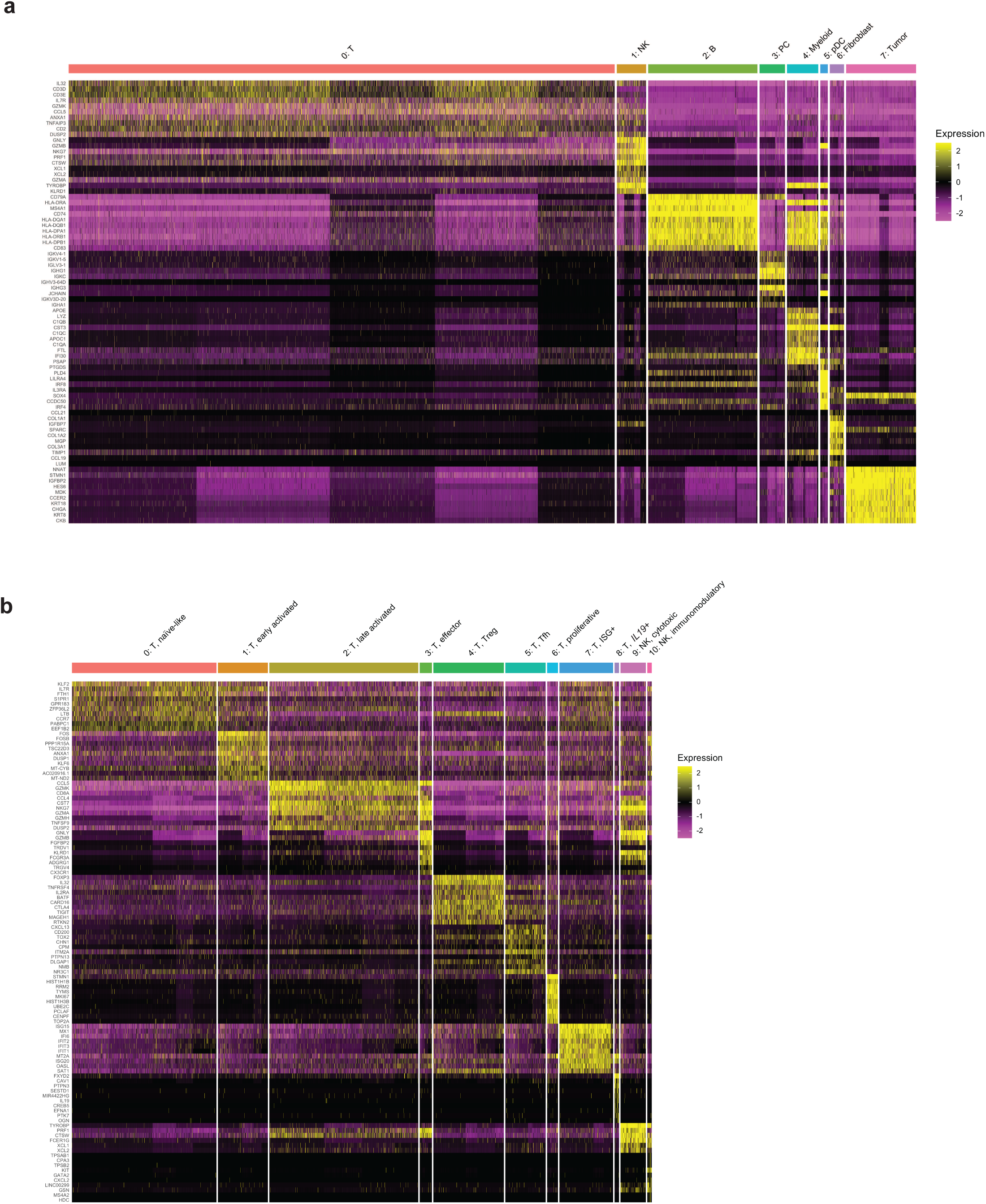
Heatmaps of marker genes for cell clusters identified in Pt 6 tumor. **a, b** Marker gene heatmaps for major cell types (a) and T/NK sub-clusters (b).

**Supplementary Figure 7.**
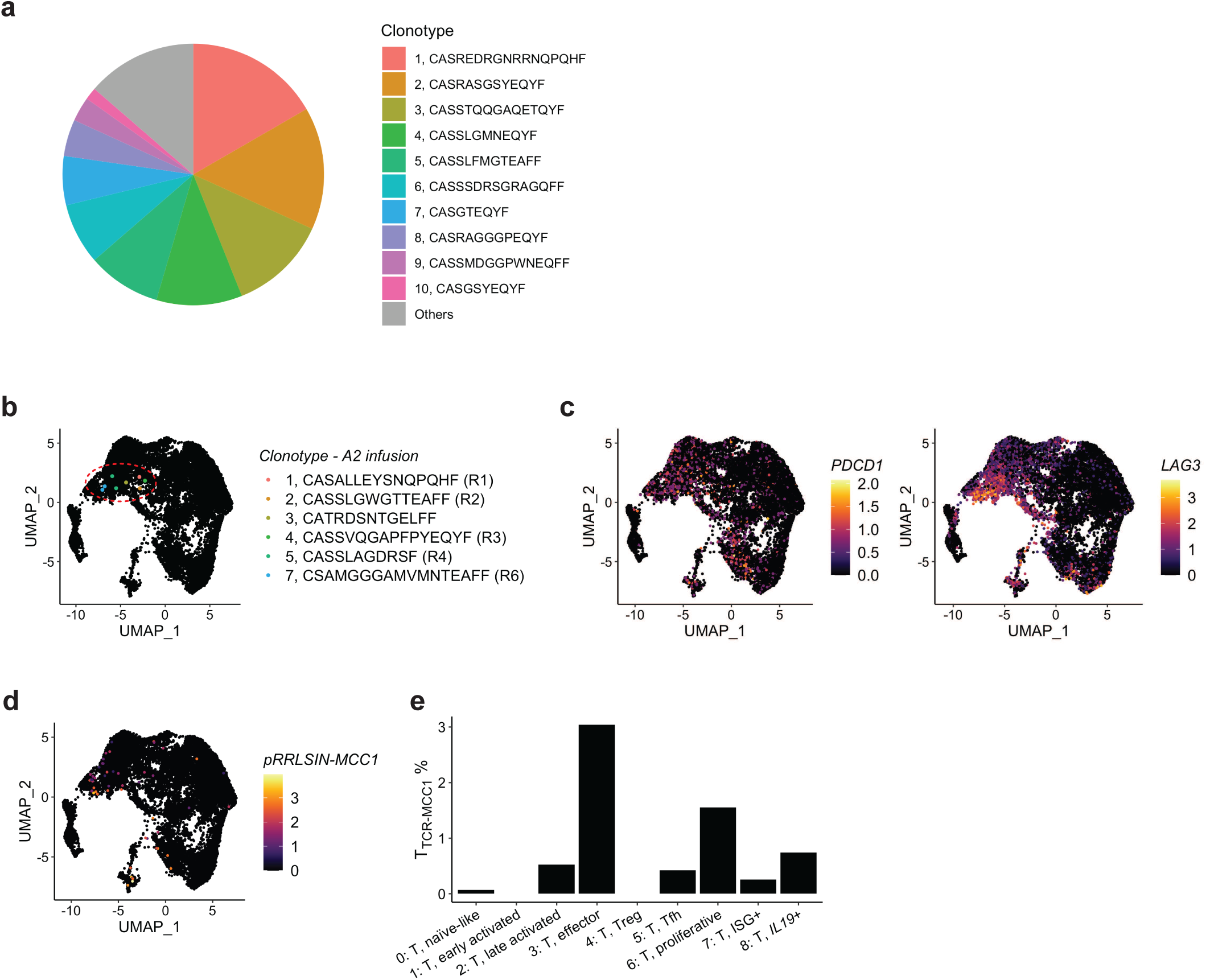
Pt 6 old infusion clonotype and T_TCRMCC1_ cells show distinct phenotypes in tumor. **a** Clonotypes in the Pt 6 old infusion product targeting sTAg_83-91_. **b** Distribution of old infusion clonotypes specific to LTAg_15-23_ projected on the UMAP. **c** Expression of *PDCD1* and *LAG3*, two marker genes of the T cell sub-cluster “2: T, late activated.” **d** TCR_MCC1_ transcript expression on the UMAP embedding. **e** Frequency of T_TCR-MCC1_ cells in T cell sub-clusters in Pt 6 tumor.

**Supplementary Table 1. Tested self peptides that partially shares amino acids with LTAg_15-23_.**

**Supplementary Table 2. HLA alleles on tested LCLs.**

**Supplementary Table 3. Biopsies.**

**Supplementary Table 4. scRNA-seq pre-processing meta data.**

**Supplementary Table 5. Marker genes of clusters identified in scRNA-seq integrated analysis of T_TCR-MCC1_ cell-containing data.**

**Supplementary Table 6. HLA LOH events.**

**Supplementary Table 7. Non-synonymous somatic SNV events in patient tumors.**

**Supplementary Table 8. CNA events in patient tumors.**

**Supplementary Table 9. CNA events in patient tumors that affected genes involved in IFN**γ **or class-I HLA presentation pathways.**

**Supplementary Table 10. IFN**γ**-downstream genes used for calculating IFN**γ **scores.**

**Supplementary Table 11. Marker genes of major cell types identified in scRNA-seq integrated analysis of Pt 6 tumor.**

**Supplementary Table 12. Marker genes of T/NK sub-clusters identified in scRNA-seq integrated analysis of Pt 6 tumor.**

**Supplementary Table 13. Differentially expressed genes in Pt 6 tumors, Day 118 vs. Pre-IFN**γ

